# Liquid-biopsy transcriptomic profiling uncovers molecular mediators of resistance to androgen receptor signaling inhibition in lethal prostate cancer

**DOI:** 10.1101/2021.11.01.21265757

**Authors:** Jiaren Zhang, Bob Zimmermann, Giuseppe Galletti, Susan Halabi, Ada Gjyrezi, Qian Yang, Santosh Gupta, Akanksha Verma, Andrea Sboner, Monika Anand, Daniel J. George, Simon G. Gregory, Seunghee Hong, Virginia Pascual, Clio P. Mavragani, Emmanuel S. Antonarakis, David M. Nanus, Scott T. Tagawa, Olivier Elemento, Andrew J. Armstrong, Paraskevi Giannakakou

**Affiliations:** Meyer Cancer Center and Division of Hematology and Oncology, Department of Medicine, Weill Cornell Medicine, New York, NY, 10065, USA; Institute for Computational Biomedicine, Weill Cornell Medicine, New York, NY, 10021, USA; Drukier Institute for Children’s Health and Department of Pediatrics, Weill Cornell Medicine, New York, NY, 10021, USA; Department of Biostatistics and Bioinformatics, Duke University, Durham, NC, Duke Cancer Institute Center for Prostate and Urologic Cancers, Duke University, Durham, NC, 27710, USA; Sidney Kimmel Comprehensive Cancer Center at Johns Hopkins, Baltimore, MD 21287, USA (current address: Masonic Cancer Center, University of Minnesota, Minneapolis, MN 55455); Department of Physiology, School of Medicine, National and Kapodistrian University of Athens, Athens, Greece; Division of Life Sciences, Department of Biotechnology, Yonsei University, Seoul, Korea

## Abstract

Androgen receptor signaling inhibitors (ARSi) are a mainstay for patients with metastatic castration-resistant prostate cancer (mCRPC). However, patient response is heterogeneous and the molecular underpinnings of ARSi resistance are not well elucidated. Here we performed transcriptome analysis of circulating tumor cells (CTCs) and peripheral blood mononuclear cells (PBMC) in the context of a prospective clinical trial of men with mCRPC treated with abiraterone (Abi) or enzalutamide (Enza). CTC RNA-sequencing identified that RB loss and enhanced E2F signaling along with BRCA loss transcriptional networks were associated with intrinsic ARSi resistance, while an inflammatory response signature was significantly associated with acquired resistance. Transcriptomic analysis of matching PBMCs identified enrichment of inflammasome gene signatures indicative of activated innate immunity at progression, with concurrent downregulation of T and NK cells. Importantly, CTC gene signatures had a significant positive association with circulating immune macroenvironment (CIME) signatures. Taken together, these data demonstrate that liquid biopsy transcriptomics can identify molecular pathways associated with clinical ARSi resistance paving the way for treatment optimization in patients with mCRPC.

## Introduction

Prostate cancer is a highly heterogeneous disease driven by androgen receptor (AR) signaling. Despite initial androgen deprivation therapy, most patients with advanced disease progress with lethal metastatic castration-resistant prostate cancer (mCRPC). It is well established that AR signaling remains active in mCRPC, largely due to residual androgen levels and activating AR aberrations, continually driving disease progression (Chen et al., 2004). To suppress AR reactivation, second-generation AR signaling inhibitors (ARSi) were developed, namely abiraterone (Abi) and enzalutamide (Enza), to inhibit *de novo* androgen biosynthesis and receptor ligand binding, respectively. Both abiraterone and enzalutamide are approved as treatments for men with mCRPC (Beer et al., 2014; de Bono et al., 2011; Ryan et al., 2013; Scher et al., 2012) and potent AR inhibitors have also demonstrated improved survival in men with metastatic hormone sensitive prostate cancer (mHSPC) where they are commonly used in current practice (Armstrong et al., 2019b; Chi et al., 2019; Davis et al., 2019; Fizazi et al., 2017; James et al., 2017).

However, the effectiveness of Abi/Enza treatment in men with mCRPC is varied and transient, and at least one quarter of patients show no evidence of clinical response, while the majority of initial responders progress within a year or two from treatment initiation (Beer et al., 2014; Scher et al., 2012). In addition, cross-resistance between AR inhibitors is common, with few patients having sustained clinical benefit from sequential AR inhibition (de Bono et al., 2018; Khalaf et al., 2019). This response profile is likely due to mechanisms of intrinsic (*de novo)* or acquired (treatment-induced) resistance. Several such mechanisms have been identified, including AR gene amplification, AR mutations (Joseph et al., 2013; Korpal et al., 2013), expression of the truncated constitutively-active AR splice variants (Antonarakis et al., 2014; Armstrong et al., 2019a; Mostaghel et al., 2011; Nadiminty et al., 2013), lineage plasticity and neuroendocrine differentiation (Brown et al., 2021b; Ku et al., 2017) to name a few (Watson et al., 2015). Nevertheless, these mechanisms do not fully explain clinical resistance phenotypes, suggesting the presence of additional, currently unidentified, mechanisms of escape.

The gap in our understanding of the molecular underpinnings of clinical ARSi response/resistance is further exacerbated by the scarcity of serial tumor biopsies, preventing us from dissecting the molecular changes underlying disease progression and response to treatment. In a recent study, transcriptomic profiling of biopsies collected before and after ARSi exposure in a small cohort of 14 men with mCRPC, implicated EMT and TGF-b pathways in ARSi resistance (He et al., 2021). Unfortunately, in this study only six samples were treatment-naïve and only four of the 14 patients had serial biopsies available, highlighting the shortage of available tissue for molecular analysis and limiting the clinical relevance of the observation. Previous work has emphasized the importance of changes in AR expression, epithelial plasticity, stemness, and TGF-beta signatures, and TP53 or RB loss as important to AR therapy resistance in this setting but phenotypic signatures have not been prospectively validated in clinical settings. (Abida et al., 2019; Annala et al., 2018; Chen et al., 2019; Wyatt et al., 2016) To overcome these limitations, we conducted a liquid biopsy-based prospective, multicenter clinical study of men with mCRPC receiving either Abi or Enza (PROPHECY)(Armstrong et al., 2019a). Circulating tumor cells (CTCs) have been repeatedly used as an alternative, validated source of tumor tissue, allowing longitudinal molecular characterization of metastatic disease (Antonarakis et al., 2017). Thus, in the context of this study, we obtained CTCs at baseline (before treatment) and at the time of progression on Abi/Enza, performed a comprehensive transcriptomic analysis of CTC and paired leukocyte RNA and correlated with clinical outcomes to identify mechanisms of ARSi resistance. We performed a transcriptomic analysis of matching peripheral blood mononuclear cells (PBMCs) in order to uncover potential involvement of the circulating immune macroenvironment (CIME) in ARSi resistance given prior studies suggesting increased myeloid inflammation and loss of T cell function with increased mortality in men with mCRPC (Olmos et al., 2012; Ross et al., 2012). Here we identified transcriptional networks indicative of RB loss, E2F signaling and BRCA loss associated with intrinsic ARSi resistance and worse overall survival. Furthermore, ARSi progression was associated with CIME alterations including enrichment of gene signatures reflective of innate immunity with concomitant depletion of T-cell and NK cell signatures. Collectively, our transcriptomic analyses represent one of the most extensive prospective characterizations of liquid biopsies in patients with mCRPC and identify novel gene expression networks associated with ARSi resistance.

## Results

### Liquid biopsy transcriptomics identify CTCs as transcriptionally distinct from PBMCs in patients with mCRPC

We present comprehensive analysis of CTC molecular profiles obtained in the context of a prospective clinical study of men with mCRPC treated with abiraterone or enzalutamide (PROPHECY), with the overall objective of developing a first-in-field CTC-based molecular taxonomy of ARSi resistance (Armstrong et al., 2019a). Prior studies from this cohort of patients have demonstrated the importance of AR-V7 detection in CTCs as well as CTC neuroendocrine phenotypes and a range of CTC DNA copy number alterations in abi/enza outcomes (Armstrong et al., 2020; Brown et al., 2021a; Gupta et al., 2021). However, the transcriptional mediators of AR therapy resistance or the immune signatures associated with outcomes were not described previously.

Liquid biopsy transcriptomic analysis was performed in the first 40 enrolled patients of the 118 total participants. Peripheral blood was collected at baseline, before ARSi treatment initiation (BS, n=40) and at the time of progression in a subset of men (PR, n=22). CTCs and matching PBMCs were isolated at each time point and subjected to RNA-sequencing (RNA-seq) (Figure 1A).

**Fig. 1.**
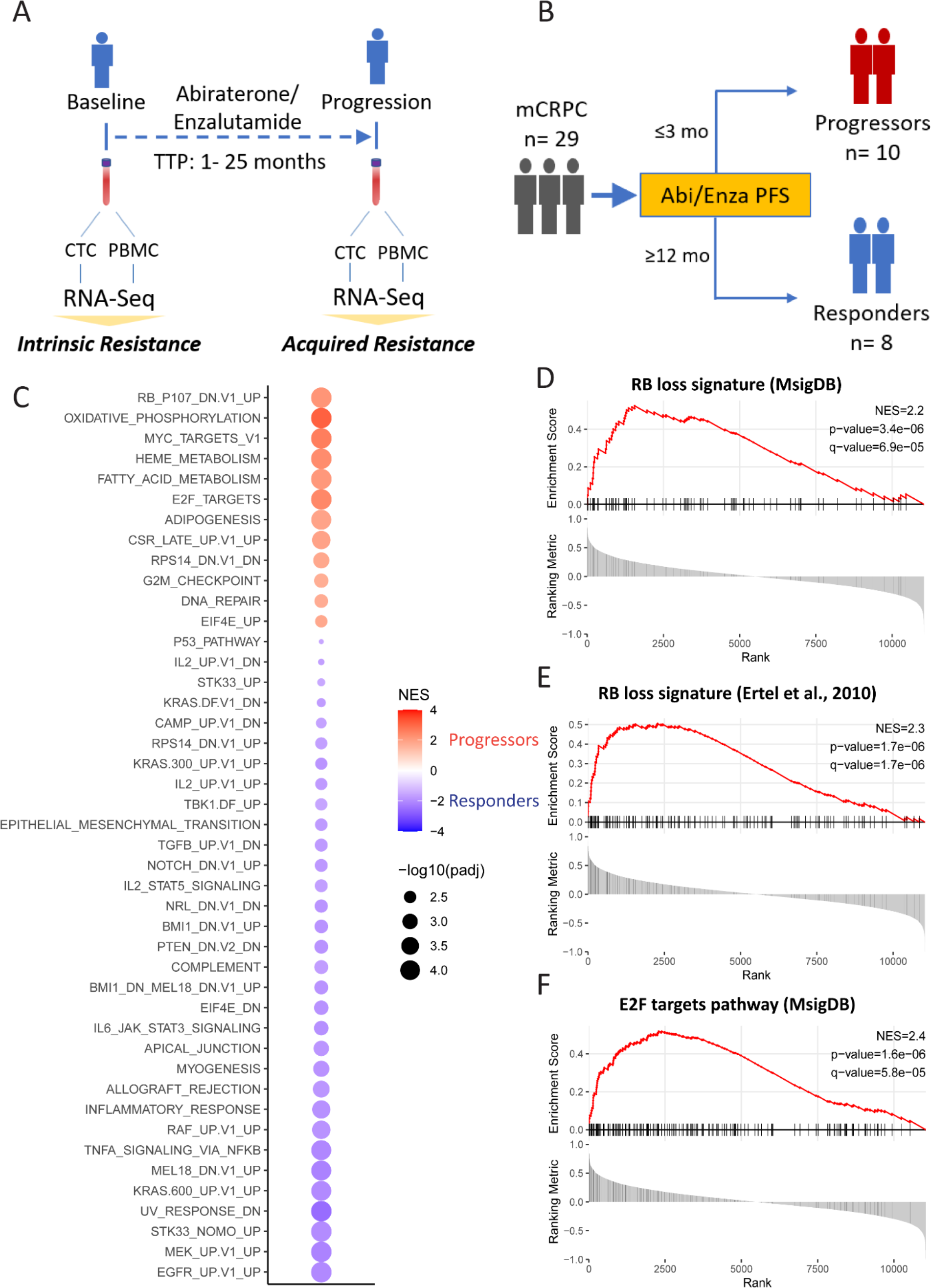
RB loss signature and E2F targets were among the most enriched pathways in progressors at baseline. **1A.** PROPHECY clinical trial high-throughput cohort sampling schema. CTCs and PBMCs were collected at baseline and progression and subjected to RNA sequencing. **1B.** Sampling of Progressors and Responders at baseline (BS). **1C.** Hallmark and Oncogenic pathways enriched in progressor CTC samples at baseline shown in red (Progressors, n=10) as compared with responders shown in purple (Responders, n=8). We detected a total of 12 hallmark and oncogenic pathways specifically enriched in ARSi progressors. Results are ranked based on adjusted p-value and color-coded based on normalized enrichment score. **1D.** GSEA enrichment plot showed RB_P107_DN.V1_UP pathway (MSigDB) significantly enriched in CTCs from progressors at BS. **1E.** GSEA enrichment plot using previously published of RB loss signature (Ertel et al 2010) shows significant enrichment in CTCs from progressors at BS. **1F.** GSEA enrichment plot showed E2F targets hallmark pathway significantly enriched in CTCs from progressors.

The resulting RNAseq datasets were subjected to a stringent filtering process based on the number of mapped reads and expression of epithelial- and prostate lineage-specific marker genes, and the absence of leucocyte transcripts, as previously described (Miyamoto et al., 2015) (Figures S1-S3). This resulted in 29 CTC-evaluable and 37 PBMC-evaluable samples at BS, and 18 CTC- and 20 PBMC-evaluable samples at PR which were used for all downstream molecular analyses. To ensure that there was no bias introduced by the filtering process, we compared the clinical baseline characteristics of the 29 patients with evaluable CTCs *vs* the overall PROPHECY cohort (Armstrong et al., 2019a) of 118 patients or the 11 patients excluded from BS analysis due to insufficient CTC RNA (Table 1). No differences were identified indicating that the filtering process did not select for a specific patient subpopulation.

**Table 1.** Clinical characteristics of patients according to CTC filtering status.

|  | **Patients with**<br>**evaluable**<br>**CTCs**<br>**(N=29)** | **Patients with**<br>**non-evaluable**<br>**CTCs**<br>**(N=11)** | **Patients with**<br>**high**<br>**throughput**<br>**(N=40)** | **Patients with**<br>**non-high**<br>**throughput**<br>**(N=78)** |
| --- | --- | --- | --- | --- |
| **Median Age in years**<br>**(range)** | **75 (48,91)** | **74 (57,85)** | **74 (48,91)** | **72 (44, 92)** |
| **Death events, no. (%)** | **26 (89.7)** | **9 (81.8)** | **35 (87.5)** | **54 (69.2)** |
| **Median OS (95% CI)** | **24.5 (18.9,**<br>**30.2)** | **19.6 (16.9, NR)** | **23.5 (18.2, 30.1)** | **19.2 (15.8, 25.0)** |
| **Median Follow-up in**<br>**Alive patients (range)** | **37.2 (32.6,**<br>**42.3)** | **32.3 (28.4, 36.2)** | **36.2 (28.4, 42.3)** | **31.0 (3.4,39.3)** |
| **CTC number, median and range** |  |  |  |  |
| **Epic (baseline)** | **1.18 (0, 283)** | **1.49 (0, 41.7)** | **1.30 (0, 283)** | **1.56 (0, 917)** |
| **Variable**<br>**All_Total_CKpos_mL** | **(1 missing**<br>**sample)** | **(2 missing**<br>**samples)** | **(3 missing**<br>**samples)** | **(8 missing**<br>**samples)** |
| **Epic (baseline)**<br>**Variable**<br>**epic_bl_traditional_ctcs_**<br>**ml** | **1.05 (0, 334)**<br>**(1 missing**<br>**sample)** | **2.20 (0, 52.1)**<br>**(2 missing**<br>**samples)** | **1.10 (0, 334)**<br>**(3 missing**<br>**samples)** | **1.50 (0, 2290)**<br>**(8 missing**<br>**samples)** |
| **Cellsearch (baseline)** | **2.50 (0, 1720)**<br>**(1 missing**<br>**sample)** | **5.00 (0, 107)** | **3.00 (0, 1720)**<br>**(1 missing**<br>**sample)** | **5.00 (0, 13000)**<br>**(7 missing**<br>**sample)** |
| **Sites of Metastases – no. (%)** |  |  |  |  |
| **Lymph Node Only** | **0 (0)** | **0 (0)** | **0 (0)** | **7 (9)** |
| **Bone Only** | **6 (20.7)** | **3 (27.3)** | **9 (22.5)** | **18 (23)** |
| **Lung Only** | **0 (0)** | **0 (0)** | **0 (0)** | **0 (0)** |
| **Multiple Sites** | **22 (75.9)** | **8 (72.7)** | **30 (75.0)** | **52 (66.7)** |
| **Prior Taxane**<br>**Chemotherapy – no. (%)** | **7 (24.1)** | **1 (9.1)** | **8 (20.0)** | **15 (19.2)** |
| **Prior Abi-Enza – no. (%)** | **9 (31.0)** | **3 (27.3)** | **12 (30.0)** | **30 (38.5)** |
| **Baseline lab values - median (range)** |  |  |  |  |
| **PSA ng/mL** | **7.53 (0.1, 319)** | **30.4 (0.3, 4190)** | **13.7 (0.1, 4190)** | **20.8 (0.2, 2020)** |
| **ALB g/L** | **3.9 (2.7, 4.4)** | **4.0 (3.2, 4.3)** | **4.0 (2.7, 4.4)** | **4.0 (2.8-4.9)** |
| **ALK U/L** | **110 (91, 117)** | **110 (91, 110)** | **110 (91, 117)** | **120 (91, 150)** |
| **HGB g/dL** | **13.2 (8.7, 15.6)** | **13.5 (9.8, 15.9)** | **13.2 (8.7, 15.9)** | **12.5 (8.8, 15.4)** |
| **LDH U/L** | **200 (100, 200)** | **200 (192, 200)** | **200 (100, 200)** | **246 (192, 618)** |
| **WBC x 10<sup>9</sup>/L** | **6.5 (4.4, 22.3)** | **5.8 (4.5, 11.0)** | **6.4 (4.4, 22.3)** | **6.1 (3.7, 9.2)** |

Next, to rule out potential leukocyte contamination in CTCs, we performed principal component analysis as well as unsupervised hierarchical clustering of CTC and PBMC samples. Both analyses indicated that CTCs are transcriptionally distinct from PBMCs (Figure S4A, B). Along these lines, gene set enrichment analysis (GSEA) revealed significant enrichment of epithelial-mesenchymal transition (EMT) pathway and AR-regulated genes (KLK2 and KLK3) in CTCs (S4D-E). Taken together, these data eliminate concerns for potential CTC misidentification or leukocyte contamination and indicate that the CTC transcriptomes represent *bona fide* PC cells.

### RB-loss and E2F target pathways are associated with intrinsic Abi/Enza resistance

To identify the molecular determinants of intrinsic (*de novo*) resistance to ARSi treatment, we examined the baseline transcriptomic profiles of mCRPC patients, comparing extreme responders or patients with progression-free survival (PFS) ≥12 months (n=8; “responders”) and extreme non-responders or patients with a PFS ≤3 months (n=10; “progressors”) (Figure 1B). We selected 12 months as a benchmark of clinical benefit in this high risk mCRPC population, given that the median PFS in this population was only 5.8 months (Armstrong et al., 2019a).

To identify pathways overrepresented in ARSi progressors at baseline, we performed gene set enrichment analysis (GSEA) using the Hallmark (n=50 pathways) and Oncogenic (n=189 pathways) gene set collection (Molecular Signatures Database, MSigDB). Using as input CTC transcriptional profiles and applying a stringent threshold for GSEA (FDR ≤0.01) we identified a total of 12 pathways, among the 239 tested, as significantly enriched in progressors relative to responders (Figure 1C). We found that the RB-loss signature (genes upregulated following RB knockout) was the most statistically significantly enriched pathway (Figure 1C). This overrepresentation of RB-loss signature in progressors (q-value=6.9x10^-5^) is depicted in the enrichment plot (Figure 1D) and was further corroborated using an independent, previously described “RB-loss signature” consisting of 158 genes identified in preclinical models of RB genomic loss and clinically validated in cancer patients (Ertel et al., 2010) (Figure 1E). Taken together, these results suggest that the RB-loss-associated transcriptional output, identified herein in patient CTCs, has a role in intrinsic ARSi resistance. As genomic loss of RB1 is one of the most frequent alterations in CRPC, our data are consistent with a recent report retrospectively correlating genomic RB1 alterations with shorter time on treatment with Abi/Enza (Abida et al., 2019). Interestingly, increased activity of the E2F target pathway was also significantly enriched in progressor CTCs at baseline as shown in the dot and enrichment plots (q-value=5.8x10^-5^) (Figure 1C, F).

It is well established that RB1 is a negative regulator of E2Fs and that loss of RB1 tumor suppressor function activates E2F signaling including target genes involved in cell cycle progression and DNA-repair (Sharma 2010) (Chen et al., 2009). Herein, we identified concurrent transcriptional outputs from RB loss and E2F targets pathways in progressor CTCs, suggesting the presence of an RB/E2F regulatory axis. To confirm this observation we generated single-sample GSEA (ssGSEA) scores for RB loss (Ertel et al., 2010) and E2F target (MSigDB) for each patient, and observed a strong correlation between the two signatures (R=0.93, p-value<0.01) indicating that the two pathways are transcriptionally and, by implication, functionally linked (Figure 2A).To ensure that this correlation is not driven by the common 56 genes shared by the two signatures (Figure 2A, Venn Diagram shaded green area), we performed a similar analysis, this time excluding the common 56 genes (Figure 2B). Our results identified a similarly high correlation (R=0.83, p-value<0.01) between RB loss/E2F target pathways, suggesting that the correlation is due to independent signaling, and driven by a mechanistic, biological link. Next, to determine the clinical significance of these two signatures in our patient cohort we examined their association with clinical outcomes and showed that patients with high RB loss or E2F signature scores at baseline experienced shorter overall survival (Figure S5).

**Fig. 2.**
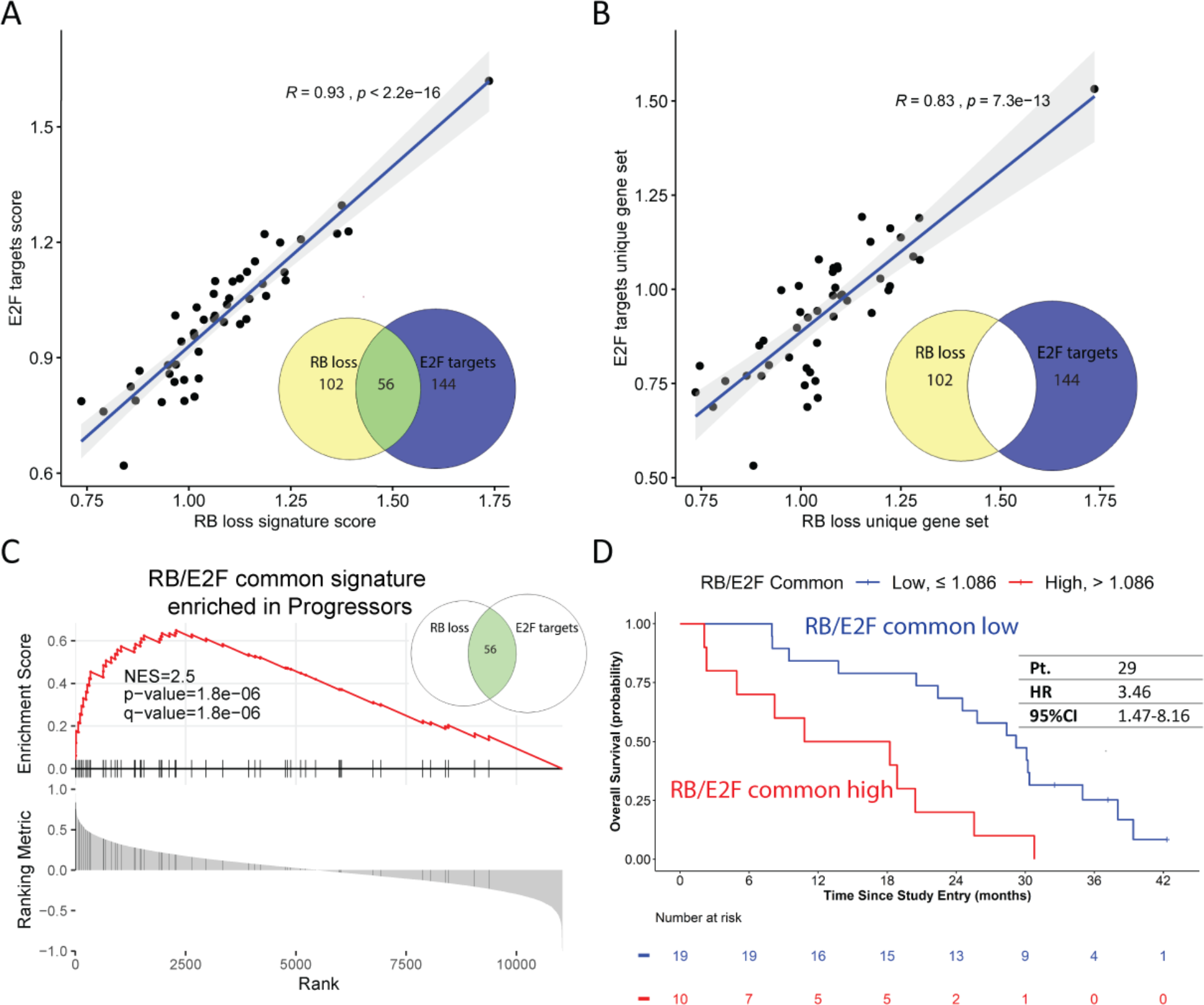
RB loss signature and E2F target gene signature are highly correlated and are associated with overall survival. Single sample enrichment score (ssGSEA) was calculated for RB-loss signature (Ertel et al., 2010) and E2F targets HALLMARK signature (MSigDB) through GSVA using genome-wide log2 TPM values as input. **2A.**E2F targets signature and RB loss signature are highly and significantly correlated in all CTC samples. **2B**. The 144 E2F target genes not-overlapping with RB-loss (E2F targets unique gene set) and the 102 RB loss signature genes, not-overlapping with E2F targets (RB loss unique gene set) remain highly and significantly correlated in CTC samples, suggesting the correlation is driven by independent signaling. **2C**. GSEA analysis using the RB_E2F common signature (56 overlapping genes), shows significant enrichment in progressors at BS. **2D**. Kaplan-Meier curves displaying the estimated survival probability for 2 groups of mCRPC patients at BS dichotomized by an optimized cutoff signature score. Overall survival by RB loss and E2F were evaluated using a univariate proportional hazards (CoxPH) model. Hazard Ratio (HR) and 95% confidence interval (CI) are shown in the table. Vertical mark in the curve indicates censored patients.

However, the large number of genes in each signature (RB loss, n=158; E2F, n=200) prompted us to generate a new condensed signature, comprised of the 56 common core genes, hereafter RB/E2F common. Enrichment analysis showed that the RB/E2F common signature was overrepresented in progressors (Figure 2C) similar to our results with each individual signature (Figure 1E, F), suggesting appropriate representation of the RB/E2F regulatory axis. Furthermore, the RB/E2F common signature was significantly associated with worse overall survival (Figure 2D), showing a higher hazard of death as compared with each individual signature (Figure S5). This RB/E2F common signature showed an association with ARSi specific PFS (HR=1.80; 95%CI=0.81-4.00).

In addition to being prognostic, RB/E2F common signature score was a viable classifier of progressors (AUC=0.75, 95% CI=0.56-0.93 on ROC analysis), suggesting that it may serve as a predictive biomarker of ARSi response which should be further investigated (Figure S6).

### A novel BRCA-loss transcriptomic signature “BRCA40” in CTCs is associated with intrinsic resistance to ARSi

Interestingly, the RB/E2F common signature contains a number of genes involved in DNA damage response and repair, including CHEK1, RAD21, MSH2 and both BRCA1 and BRCA2. Along these lines, our analysis identified enrichment of DNA repair pathway in progressor CTCs at baseline concomitantly with RB-loss and E2F targets (Figure 1C). It is well established that genes implicated in DNA damage response and repair (DDR) machinery are frequently co-altered in mCRPC, and genomic deletion or inactivating mutations of BRCA1 and BRCA2 are found in up to 10-15% of patients with mCRPC (Abida et al., 2019). Thus, we set out to identify a gene set whose expression phenocopies the BRCA loss-of-function phenotype. We started with gene sets previously associated with higher patient risk for breast cancer using integrated network modeling strategies centered on BRCA, described in Pujana et al. (Pujana et al., 2007). Enrichment analysis identified 4 of the 6 BRCA-related gene sets overrepresented in progressor CTCs at baseline (Figure 3A). Interestingly, no BRCA-loss signatures were enriched in responder CTCs. Given that these BRCA-gene sets were only validated in microarray data from breast tissue, we sought to establish a BRCA-loss signature applicable to patients with mCRPC. Additionally, given the large size of these gene sets (ranging from 57 to 1627 genes), we derived a new more concise gene set comprising the 40 overlapping core genes, hereafter referred to as “BRCA40”, shared among all enriched BRCA-gene sets in our cohort (Figure S7A).

**Fig. 3.**
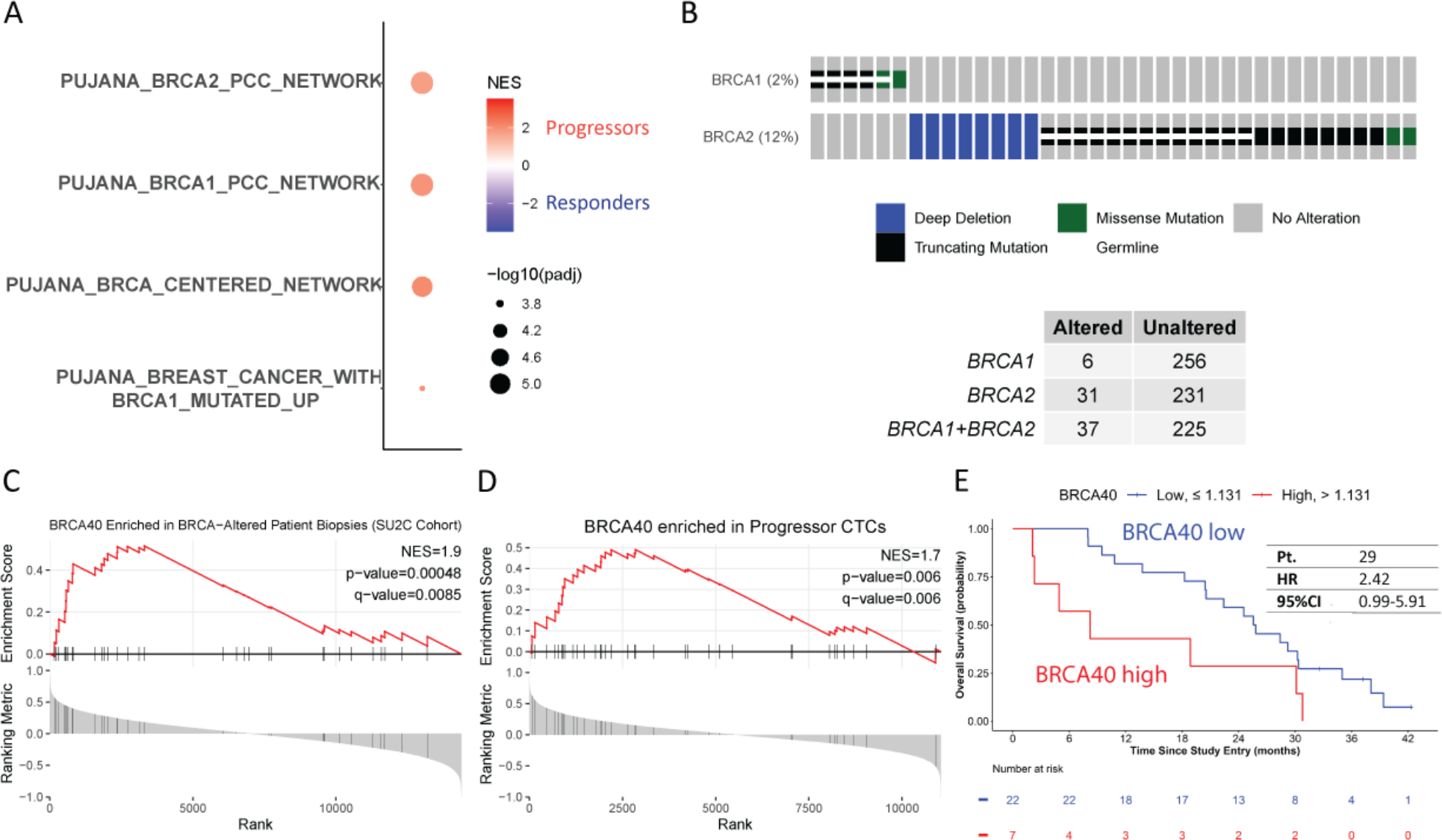
BRCA loss transcriptional gene signatures were significantly enriched in ARSi progressors at baseline. **3A**. We detected 4 BRCA-loss related gene expression pathways (Pujana et al., 2007) only enriched in progressors with FDR threshold ≤ 0.01. Results are ranked based on adjusted p-value and color-coded based on normalized enrichment score. **3B.** Oncoprint summary of inactivating genomic BRCA alterations (missense/truncating mutation and depletion) in mCRPC patients from the SU2C database. BRCA alterations were observed in 37 (of 262; 14%) patients with mCRPC. Samples with white stripes through indicate germline mutation. **3C.** BRCA40 transcriptomic signature was significantly enriched in BRCA-gene-genomic-altered mCRPC patients as compared to unaltered patients from SU2C database. **3D**. BRCA40 gene signature was significantly enriched in CTCs from progressors at baseline (BS) **3E.** Kaplan-Meier curves displaying the estimated survival probability for 2 groups of patients with mCRPC at baseline (BS), dichotomized by an optimized cutoff ssGSEA score of BRCA40 signature using the Lau92 p-value approximation method using limiting distribution by a Brownian bridge (Lausen and Schumacher, 1992). Overall survival by BRCA40 was evaluated using a univariate proportional hazards (CoxPH) model. Hazard ratio (HR) and 95% confidence interval (CI) were shown in the table. Vertical mark in the curve indicates censored patients.

To examine whether the BRCA40 signature reflects BRCA genomic alterations we tested it in the publicly available integrative database of SU2C mCRPC patient cohort (Abida et al., 2019). Using paired expression and genomic alteration data from 262 patients, 37 (14%) of which had BRCA alterations (deletions or mutations) (Figure 3B), we found that the BRCA40 signature was significantly enriched in the BRCA-altered SU2C cohort when compared to unaltered patients (Figure 3C). Additionally, we found that the BRCA40 signature performed similarly (q-value=8.5x10^-4^) to the top enriched BRCA-related gene set (BRCA2 PCC network, n=423 genes, q-value=7.1x10^-4^) in identifying BRCA-altered SU2C patients (Figure S7B-G). Applying this novel BRCA40 signature in our patient cohort we found it was significantly enriched in baseline progressors CTCs (Figure 3D), and was highly correlated with RB loss, E2F target, and RB/E2F common signatures (Figure S7H- J), also identified as enriched in ARSi-resistant patients. In addition, using BRCA40 ssGSEA scores we observed that patients with high BRCA40 scores experienced shorter overall survival (Figure 3E). In addition to being prognostic, this BRCA40 signature also showed an association with ARSi specific PFS (HR=1.68; 95%CI=0.68-4.14) Taken together, these data identify BRCA40 as the first BRCA loss-of-function transcriptomic signature in mCRPC and suggest a potential mechanistic link between BRCA loss and RB/E2F axis in ARSi resistance.

### Gene signatures are enriched in CTCs at Progression (acquired ARSI resistance CTCs)

In addition to identifying genes and pathways associated with *de novo* (intrinsic) resistance to ASRi treatment, we sought to investigate potential mechanisms contributing to treatment-induced (acquired) resistance (Figure 1A). In order to accomplish this goal, we compared the CTC transcriptomic profiles obtained from patients at progression (n=18, median PFS = 3.8 months, 95% CI= 2.8-9.5 months) with those obtained at baseline (n=29). Differential gene expression analysis followed by GSEA identified 14 pathways significantly enriched at progression (PR) relative to baseline (BS) (Figure 4). We observed enrichment for the inflammatory humoral response at progression (PR), evidenced by the top enriched pathways being inflammatory response, complement and enrichment of TNF-α signaling via NF-DD pathway (Figure 4A, B).

**Fig. 4.**
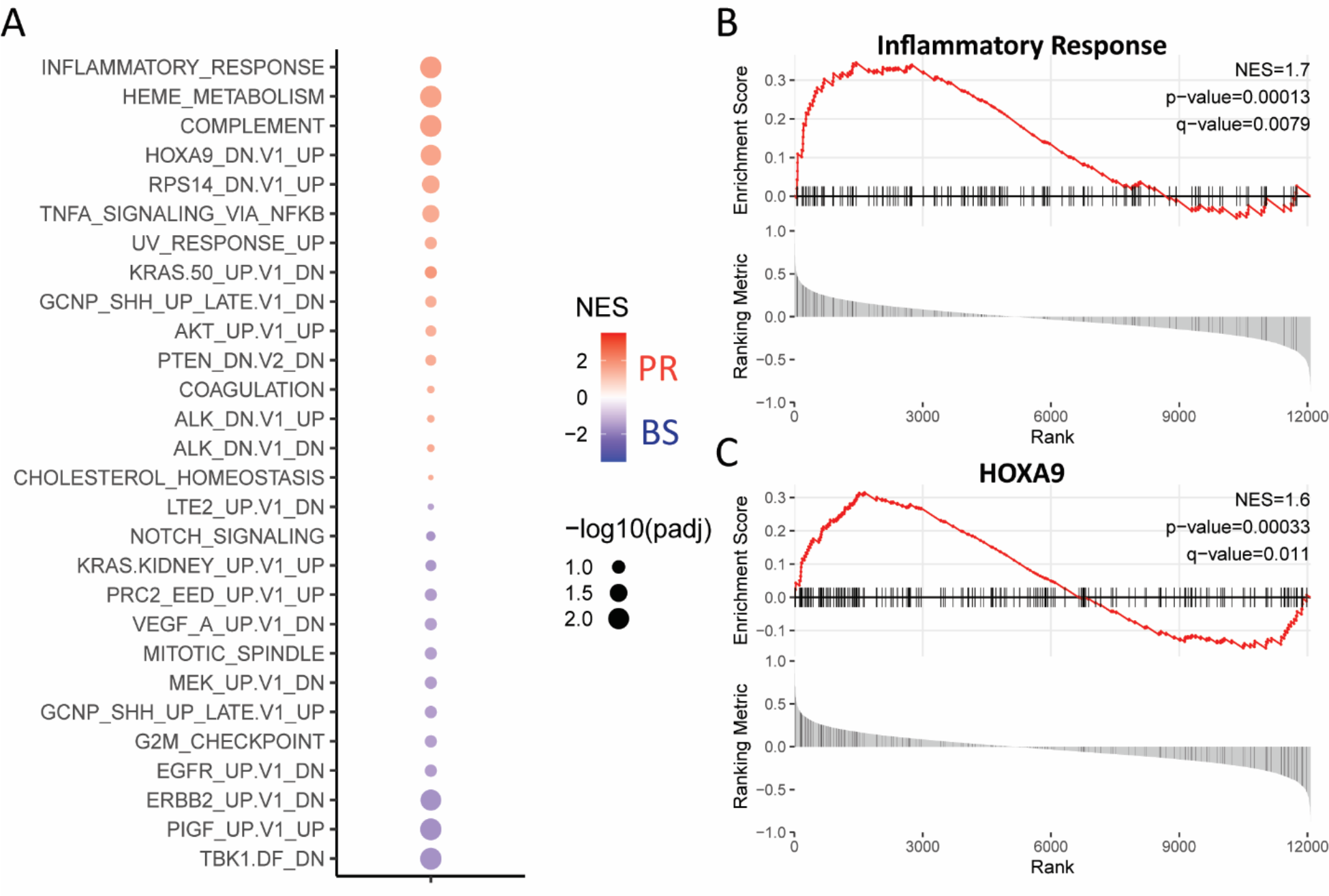
Pathways associated with acquired Abi/Enza resistance. **4A.** Hallmark and Oncogenic from MSigDB pathways enriched in progression (PR) CTCs as compared to baseline (BS) CTCs. Adjusted p-value cutoff is 0.25. **4B.** The inflammatory hallmark pathway was significantly enriched in PR CTCs in comparison with BS CTCs. **4C.** HOXA9 pathway was significantly enriched in PR CTCs in comparison with BS CTCs.

Heme metabolism was the second most enriched pathway in CTCs at progression. Intriguingly, leading edge genes analysis identified key regulators of iron metabolism (SCNA, HMBS and TFRC; data not shown), which are frequently upregulated in cancer cells (Shekoohi et al., 2021). Considering that Heme metabolism pathway is found enriched exclusively in the CTC compartment and that the heme structure is an essential cofactor for enzymatic complexes of the electron transport chain and the cytochromes, we posit that this enrichment reflects augmented tumor-specific metabolic requirements at progression.

Interestingly, we identified HOXA9 gene inactivation signature significantly enriched in patient CTCs at progression (Figure 4A, C). HOXA9 functions as a tumor-suppressor gene in breast cancer, transcriptionally activating PTEN through promoter binding. Thus, HOXA9 silencing in breast cancer preclinical models was shown to suppress PTEN expression leading to the activation of the PI3K-AKT signaling pathway (Mouw et al., 2014). In agreement with these mechanistic studies, our CTC transcriptomic analyses identified enrichment of PTEN loss and AKT activation signatures concomitantly with HOXA9 inactivation at progression (Figure 4A, C).

Taken together, these findings indicate a potential mechanistic link between HOXA9 and PTEN/AKT axes in patients with mCRPC and provide further support for ongoing clinical trials combining AKT inhibitors with ARSi in the context of PTEN loss (Sweeny et al 2021 PMID: 34246347)

### Circulating immune macroenvironment transcriptional analysis identifies coordinated alterations and crosstalk with CTCs at progression

The significant enrichment of pathways associated with inflammation and immune pathway response in CTCs at progression prompted us to investigate the coordinated alterations in the circulating immune macroenvironment (CIME) in our patient cohort using patient and time-matched blood samples.

First, we analyzed the CIME transcriptomic profiles obtained from patients at BS (n=37) and PR (n=20) using the xCell platform (Aran et al., 2017). We observed significant up- regulation of the pan-macrophage signature with concomitant depletion of CD8+ T-cells signature at PR (Figure S8). To expand on this analysis, we analyzed CIME transcriptomes using a framework of 63 annotated blood-derived transcriptional modules formed by genes coordinately expressed across multiple disease data sets (Banchereau et al., 2016; Chaussabel et al., 2008). We identified a significant enrichment in myeloid-lineage, neutrophil and inflammation gene modules at progression. Concurrently, we observed significant downregulation of CD8+ (and to a lesser extent CD4+) T-Cell and cytotoxic NK-cell modules (Figure 5A, Figure S9). These results, which corroborate and expand the xCell analysis, were independently validated by GSEA (Figure S10).

**Fig. 5.**
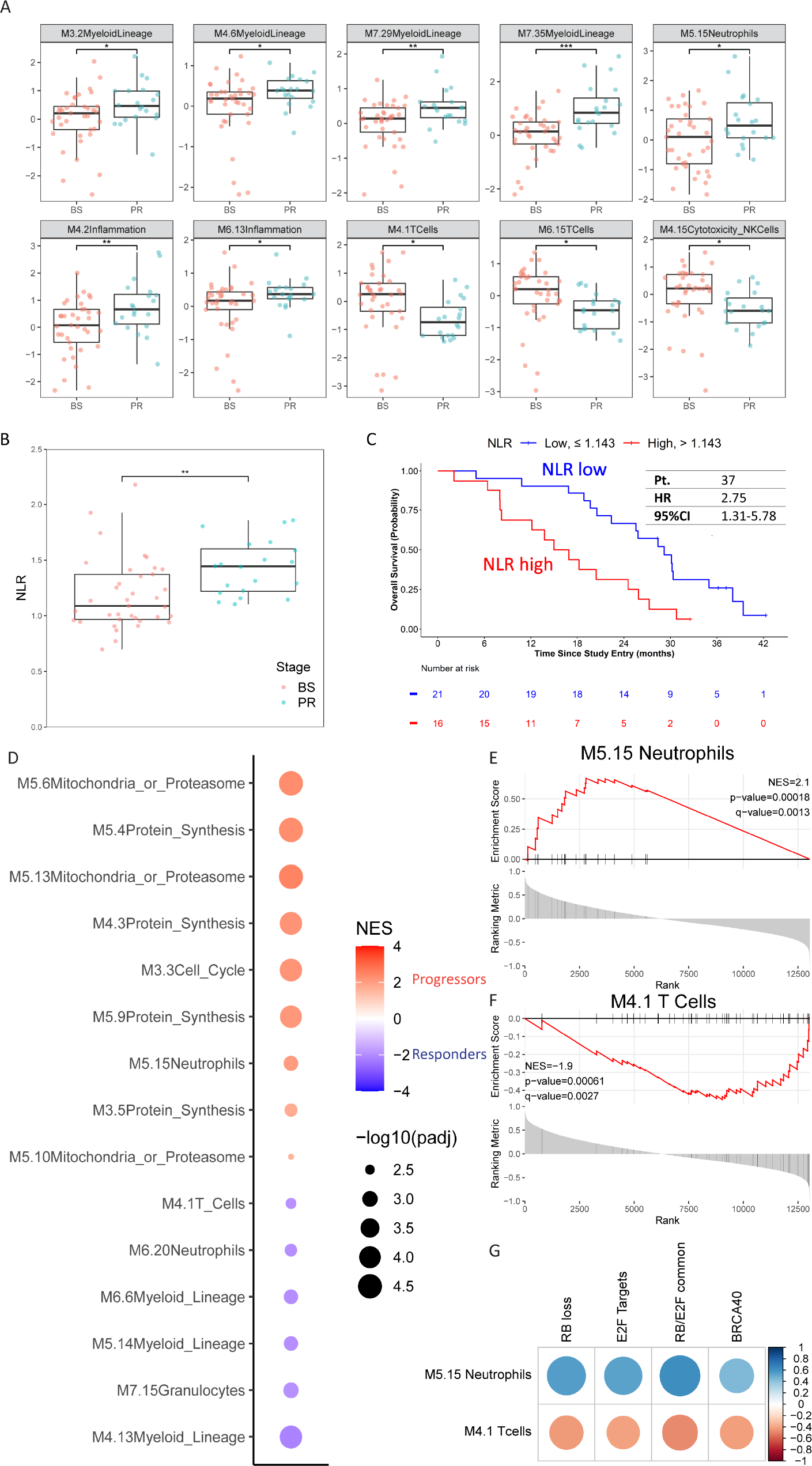
Circulating immune macroenvironment (CIME) transcriptomic analysis identified enrichment of myeloid lineage modules with concurrent depletion of lymphoid lineage modules at ARSi progression, and a correlation with CTC-derived pathways of intrinsic resistance at baseline. 5A. Myeloid lineage transcript modules (top row, first 4 modules), neutrophils and two inflammation modules are up-regulated at progression (PR) as compared to baseline (BS) with concomitant down regulation of T-cells (M4.1 transcripts are expressed in both CD4+ and CD8+ cells while M6.15 transcripts are expressed exclusively in CD8+ T cells) and Cytotoxicity NK cells transcript modules. Boxplots of the 10 significantly differentially expressed immune modules from the comparison of PR PBMCs with BS PBMCs. Adjusted p-value cutoff is 0.05. 5B. Transcriptomic Neutrophil to Lymphocyte Ratio (NLR) was significantly elevated at progression. 5C. Kaplan-Meier curves displaying the overall survival probability for 2 groups of NLR mCRPC patients at BS dichotomized by an optimized NLR value. Overall survival was evaluated using a univariate proportional hazards (CoxPH) model. Hazard ratio (HR) and 95% confidence interval (CI) were shown in the table. Vertical mark in the curve indicates censored patients. 5D. Dot plot of enriched CIME modules at baseline, using the 63 functionally annotated gene sets (Chaussabel et al., 2008; Banchereau et al., 2016) as a customized database for GSEA; p-value cutoff is 0.01. Note the enrichment of the M5.15 Neutrophil module (5E) and the depletion of the M4.1 T cell module in Progressors (5F). 5G Correlation matrix plot between CIME and CTC transcriptomic pathways at baseline. The M5.15 Neutrophil module shows positive correlation with the CTC derived RB loss, E2F targets, RB/E2F common and BRCA40 core gene signatures enriched in patients with intrinsic ARSi resistance, while the CIME M4.1 T cell module is negatively correlated with these CTC signatures at baseline. Matrix plot color intensity and circle size are proportional to the correlation coefficient (R value).

The enrichment of the neutrophil module together with the downregulation of the T-cell modules prompted us to generate a transcriptomic-based neutrophil-to-lymphocyte ratio (tr-NLR) score. Increased NLR, based on standard neutrophil and lymphocyte peripheral blood counts, has been associated with worse clinical outcomes in patients with mCRPC (Guan et al., 2020; Mehra et al., 2017) . Herein, we identified that tr-NLR was significantly elevated at PR, likely reflecting the changing landscape of CIME and its contribution to disease progression (Figure 5B). Baseline GSEA analysis of CIME transcripts identified the same modules used in our tr-NLR calculation, as enriched (M5.15 Neutrophil) or depleted (M4.1 T cell) in progressors, suggesting a role of the CIME in intrinsic ARSi resistance (Figures 5D-E). Furthermore, high tr-NLR at baseline was significantly associated with shorter overall survival (OS) (Figure 5C), consistent with the reported prognostic value of cell count based NLR.

It is well established that the tumor microenvironment in PC plays an integral role in disease progression via crosstalk with the tumor compartment. This study provided us with the unique opportunity to test a potential link between the tumor cells and tumor microenvironment in the circulation using liquid biopsies. Thus, we set out to identify interactions between the patients’ CIME and CTC compartments and their potential role in ARSi resistance. Indeed, we observed a significant correlation between the CIME Neutrophil (positive) and T-cells (negative) modules and the CTC pathways associated with intrinsic ARSi resistance, namely RB loss, E2F targets, RB/E2F common and BRCA40 signatures (Figure 5G). Taken together, these data provide support for the presence of crosstalk between CTCs and CIME and its potential association with ARSi resistance; while suggesting the potential therapeutic benefit of immune system modulation in combination with ARSi for patients with mCRPC.

## Discussion

The vast majority of men with metastatic hormone sensitive and castration resistant prostate cancer benefit from the receipt of potent inhibitors of the androgen receptor signaling axis, resulting in improved survival and more durable remissions as compared to androgen deprivation therapy alone. Second-generation androgen receptor signaling inhibitors (ARSi), including abiraterone and enzalutamide, have induced a paradigm-shift in the management of advanced castration resistant prostate cancer (mCRPC) (Beer et al., 2014; Ryan et al., 2013). The unprecedented clinical impact of ARSi in mCRPC prompted their clinical use for the treatment of earlier stages of the disease (non-metastatic CRPC; nmCRPC) (Hussain et al., 2018) and of metastatic hormone-sensitive PC (mHSPC) (Davis et al., 2019; Fizazi et al., 2017; James et al., 2017).

Despite ARSi clinical success, there remains significant heterogeneity in the durability and robustness of response to these therapies. Critical to understanding this patient and tumor heterogeneity, as well as the emergence of primary, secondary, and cross-resistance between AR inhibitors, is the development of biomarkers to measure disease biology and connect this to the patient experience. Importantly, the molecular underpinnings of ARSi clinical resistance are incompletely understood. In this present study, we have provided clear evidence supporting both tumor and host mechanisms of resistance that contribute to AR therapy outcomes using a novel non-invasive liquid biopsy transcriptomic analysis.

Liquid biopsies have been utilized as a minimally invasive approach to investigate molecular alterations associated with ARSi resistance in patients with mCRPC (Annala et al., 2018; Casanova-Salas et al., 2021). However, most studies have used ctDNA analyses to describe the genomic landscape of mCRPC focusing mostly on the AR gene and its treatment-associated genomic alterations (Annala et al., 2021; Romanel et al., 2015). In addition, our prior work has strongly associated the detection of AR-V7 in CTCs with AR therapy resistance, and CTC phenotypes and genotypes with these outcomes (Armstrong et al., 2019a; Brown et al., 2021a; Gupta et al., 2021). However, many DNA based alterations may lack a functional impact or phenotype, illustrating the need for broader phenotypic assays related to RNA expression both of tumor and immune cell subsets, which may better capture patient outcomes.

In this study, we sought to elucidate the molecular underpinnings of clinical ARSi resistance by performing comprehensive transcriptomic profiling of CTCs and circulating immune cells derived from patients with mCRPC before the initiation of ARSi treatment and at disease progression, in the context of the prospective multicenter PROPHECY study (Armstrong et al., 2019a). Our pre-specified transcriptomic analysis of baseline CTCs nominated the RB loss signature as the top enriched pathway in patients progressing within the first three months of Abi/Enza treatment (extreme progressors), suggesting a clinical association between RB loss transcriptional output and intrinsic ARSi resistance. In agreement with our findings, genomic loss of RB1 was associated with shorter time on treatment with ARSi in patients with mCRPC in the SU2C cohort (Abida et al., 2019). RB1 is a tumor suppressor gene that is emerging as one of the most commonly altered genes in mCRPC (Abida et al., 2019; Robinson et al., 2015a). Mechanistically, it is well established that RB1 negatively regulates E2F1 activity while genomic RB1 loss leads to E2F1 cistrome expansion, and downstream cell cycle progression and cell proliferation (Dyson, 2016; McNair et al., 2018). Interestingly, recent studies using prostate cancer cell lines resistant to Abi or Enza demonstrated E2F signaling activation upon AR inhibition; however, unlike our findings, the E2F signature was not associated with RB1 downregulation (Handle et al., 2019; Kim et al., 2021). Herein, our data establish a strong association between RB loss and E2F transcriptional networks in clinical ARSi resistance, while our newly identified RB/E2F common signature is a viable classifier of ARSi progressors suggesting that it could be used as an indicator of ARSi response in future clinical studies (Chen et al., 2019). Notably, the RB1 loss transcriptomic classification identified high RB-loss even in patients with genomically intact RB (Table S1), suggesting that transcriptomic RB-loss identified an expanded pool of patients with impaired RB signaling who would be missed by genomic analysis alone.

Genomic loss of RB1 and BRCA2, located in proximity to one another on chromosome 13q, are among the most frequent genomic alterations in mCRPC. Interestingly, a recent study revealed that genomic co-loss of both RB1 and BRCA2 identifies a distinct subset of prostate cancer patients characterized by more aggressive disease and poor clinical outcomes (Chakraborty et al., 2020; Mandigo and Knudsen, 2020). In this study, we show that the transcriptional outputs from BRCA-loss (BRCA40), RB-loss and E2F targets are highly and significantly correlated with each other in CTCs, as well as associated with intrinsic ARSi resistance. These transcriptional data are consistent with the RB1, BRCA2 genomic co-loss in aggressive PC, while they reveal a previously unrecognized role for RB/E2F/BRCA axis in clinical response to standard-of-care treatment with Abi/Enza in mCRPC.

Currently, BRCA loss-of-function is determined exclusively at the genomic level (Pritchard et al., 2016; Robinson et al., 2015b). However, recent conflicting studies regarding the association between genomic BRCA1/2 loss and clinical response to ARSi treatment (Annala et al., 2018; Antonarakis et al., 2018), suggest that genomic assessment of BRCA status may not recapitulate the entire spectrum of BRCA-loss tumor phenotypes. It is well established that BRCA functional output can be determined by altered transcriptional regulation of BRCA genes via various post-genomic epigenetic mechanisms such as promoter methylation and histone acetylation (Lord and Ashworth, 2013; Pao et al., 2000; Ruscito et al., 2014). Thus, we posited that a transcriptional BRCA-loss signature would more faithfully recapitulate BRCA loss tumor phenotypes, that extend beyond genomic alterations. To fill this gap, we developed a novel BRCA-loss transcriptomic signature (BRCA40), inspired by the BRCA-centered integrated network modeling analysis performed in breast cancer (Pujana et al., 2007). Our BRCA40 signature accurately recapitulated the BRCA genomic alteration status in the SU2C mCRPC patient cohort and was significantly associated with intrinsic ARSi resistance and worse overall survival in our cohort. Similar to what we observed with RB1 loss, our transcriptomic classification (BRCA40 score) identified patients with high BRCA score (indicative of BRCA loss of function) even in patients with genomically unaltered BRCA 1 and 2 genes (Table S1). This observation suggests that our transcriptomic definition of BRCA loss could broaden the identification of patients harboring the BRCA loss phenotype.

As genomic BRCA loss has been clinically associated with sensitivity to treatment with PARP inhibitors (PARPi) in patients with mCRPC (Fong et al., 2009), we propose that our BRCA40 signature may expand and more optimally refine the patient population with functional loss of BRCA, thus, identifying patients likely to benefit from PARPi therapies, beyond those with genomic alterations. This hypothesis should be validated prospectively.

CTC transcriptomic analyses further showed enrichment in inflammatory humoral responses, upon disease progression suggesting a role for tumor derived inflammation and complement activation in treatment-induced ARSi resistance. Preclinical evidence shows that inflammation promotes prostate cancer cell survival and metastatic dissemination (Huang et al., 2001; Nguyen et al., 2014; Shukla and Gupta, 2004); while in the SU2C cohort, inflammation and complement pathways were found enriched in patients exhibiting poor response to enzalutamide treatment (Alumkal et al., 2020). While it is not yet clear whether tumor inflammatory signaling through NF-DB or TNF-α or other pathways promotes clinical resistance to AR-targeted therapies (ADT and ARSi), prior mechanistic studies have suggested an important role for NF-DB signaling in mediating AR therapy resistance (Jin et al., 2015; Liang et al., 2021; Nadiminty et al., 2015; Nadiminty et al., 2013) and further clinical investigation is warranted to determine potential synergistic effect between therapies targeting these pathways and ARSi. Furthermore, our results showing concurrent enrichment for HOXA9 gene inactivation, PTEN loss and AKT activation at progression, suggest a potential new mechanistic link between HOXA9 and PTEN/AKT pathways which may be useful in the selection of patients more likely to benefit from the combination of AKT inhibitors with ARSi in the context of PTEN loss (de Bono et al., 2019; Sweeney et al., 2021).

The profound enrichment of inflammatory pathways in patient CTCs at progression, together with the established role of inflammation in the tumor microenvironment in PC progression (de Bono et al., 2020), led us to expand our transcriptomic analysis to patient circulating immune macroenvironment (CIME). Notably, it was recently shown in patients with different tumor types that expansion of T cell clones within the tumor was paralleled by their expansion within the peripheral blood, and that patients with such expansion derived clinical benefit from with anti-PD1 treatment. These data show that blood-based analyses may mirror tumor and tumor immune microenvironment alterations and highlight the potential of liquid biopsies in predicting treatment response (Wu et al., 2020).

Transcriptomic analysis of patient CIME at progression showed downregulation of cytotoxicity NK and T cell modules, indicating suppression in adaptive immunity. Concurrently, we observed enrichment in myeloid lineage, neutrophil, and inflammation gene modules, indicating stimulation of innate immunity. These data are supported by prior cell free RNA expression signatures from whole blood in men with mCRPC, demonstrating the prognostic importance of myeloid and T cell subsets in this setting (Olmos et al., 2012; Ross et al., 2012). Central to innate immunity is the activation of the “inflammasome”, a multi-protein network driving a cascade of proinflammatory cytokines, such as IL-1 and IL- 18 (Zheng et al., 2020); while upregulation of IL-1α and IL-1β has been shown to promote tumor invasiveness, angiogenesis and metastasis (Lewis et al., 2006; Litmanovich et al., 2018; Voronov et al., 2003). In our analysis, both the inflammation (M4.2) and myeloid modules (M3.2, M4.6, M7.29 and M7.35) were enriched at progression, including genes encoding key members of the inflammasome complex, such as IL1R1, IL1RAP, IL1RN, IL1RAP, IL18R1, and IL18RAP, suggesting inflammasome activation in our patient cohort at progression. These pathways were particularly pronounced in men with CTCs harboring RB1 loss/E2F/DNA damage signatures, suggesting cross talk between tumor cells and the immune macroenvironment in promoting disease progression and aggressiveness. Interestingly, a recent study in melanoma showed that activation of inflammasome/IL1 signaling led to a reduction of cytotoxic T cells, which is also consistent with our findings (Tengesdal et al., 2021).

Taken together, the CIME transcriptional profiling identified molecular alterations at progression consistent with upregulation of innate and suppression of adaptive immunity, implicating inflammasome activation. Given that pharmacologic inhibitors of inflammasome/IL-1 signaling are already used for the treatment of autoinflammatory syndromes, our data suggest that their combination with ARSi treatment could represent a synergistic therapeutic strategy for patients with mCRPC. This remains to be confirmed clinically.

The enrichment of inflammation signaling in both CTC and CIME at progression suggests a previously unrecognized interplay between tumor cells and the immune macroenvironment, in the host’s circulation. This is further supported by the significant correlation between the CTC-derived RB/E2F and BRCA loss pathways and the CIME-based Neutrophil and T cell modules, in extreme progressors at baseline. Interestingly, a mechanistic link between RB1, E2F and T cells has been recently demonstrated in zebrafish where RB-deficient or E2F1-overexpressing fish exhibited a significant reduction in T cells during development, (Zhang et al., 2018), further supporting the presence of cross-talk between the CTC and CIME compartments.

In conclusion, these results demonstrate that transcriptomic analyses of patient liquid biopsies can identify molecular determinants of response to ARSi treatment and can be used to monitor changes in CTCs and the CIME that may have therapeutic implications. We posit that the clinical relevance of our findings could benefit mCRPC as well as mHSPC and nmCRPC, where ARSi are extensively used. We propose to validate these peripheral transcriptomic signatures in future prospective clinical trials and nominate coordinate RB loss and DNA repair deficiencies as critical determinants of patient outcomes worthy of therapeutic targeting.

## Materials and methods

### Patient characteristics and clinical study design

Men with progressive, high-risk metastatic castration-resistant prostate cancer (mCRPC) were enrolled in the multicenter, prospective, single-arm PROPHECY study (NCT02269982) (Armstrong et al., 2019a); this study evaluated the prognostic significance of biomarkers assessed in circulating tumor cells (CTCs) isolated from patients initiating treatment with abiraterone or enzalutamide. The first consecutive 40 subjects of the study enrolled at two clinical sites (Duke University and Weill Cornell Medicine) were included in the CTC-based transcriptomic analysis cohort. All patients provided written informed consent in accordance with Declaration of Helsinki ethical guidelines; Institutional Review Board (IRB) approval was obtained from Duke University IRB or Weill Cornell Medicine IRB.

The major clinical characteristics of our patient cohort are summarized in Table 1. Treatment with abiraterone acetate with prednisone or enzalutamide was continued until disease progression, unacceptable toxicities, withdrawal of the consent or death. Progression-free survival (PFS) was defined as the time from date of registration to clinical/radiographic progression or death, whichever occurred first. Overall survival (OS) was defined as the time from date of registration until death of any cause.

### Blood collection and CTC enrichment

Peripheral blood samples for CTC isolation were collected in EDTA tubes (BD Vacutainer) at prespecified time points: baseline (before treatment initiation) and at the time of disease progression. The CTC processing was based on our established Standard Operating Protocol, as previously described (Antonarakis et al., 2017; Worroll et al., 2019). Briefly, 20 mL of peripheral blood were collected from each patient at each time point; CTCs were enriched by CD45 depletion (RosetteSep Human CD45 Depletion Cocktail; Stemcell Technologies) according to the manufacturer’s instructions. Matching peripheral blood mononuclear cells (PBMCs) were also isolated from each patient at each time point, by density gradient centrifugation. The enriched CTCs and PBMCs were subsequently lysed in RLT Plus buffer (Qiagen) followed by RNA extraction for RNA-sequencing library preparation.

### RNA extraction and RNA sequencing

Liquid biopsy transcriptomic analysis was performed in the first 40 enrolled patients at Duke University and Weill Cornell Medical College, of the 118 total participants across 5 sites. These patients were consecutively enrolled at these sites as part of a prospectively defined high-throughput subcohort of the overall PROPHECY study. Peripheral blood was collected at baseline, before ARSi treatment initiation (BS, n=40) and at the time of progression in a subset of men (PR, n=22). CTCs and matching PBMCs were isolated at each time point and subjected to RNA sequencing Total RNA was extracted from the enriched CTCs and matching PBMCs using the RNAeasy Plus Micro kit (Cat. # 74034, Qiagen) as per manufacturer’s instructions. Full-length mRNA-seq libraries were generated using SMARTer Ultra Low RNA Kit (Version 4, Cat. # 634891, Takara Bio/Clontech Laboratory) (Ramskold et al., 2012). Double-stranded cDNAs were converted into sequencing ready libraries using Nextera XT DNA Sample Preparation Kit (Cat. # FC-131- 1096, Illumina). The quality control and quantity estimates for each library were done after the last purification using a hsDNA chip for BioAnalyzer (Agilent). After normalization, the libraries were multiplexed and sequenced as 8 samples/lane in an Illumina HiSeq4000 machine with configuration of paired-end 2x50 bp.

### RNA-Seq data processing and sample quality control

RNA-Seq raw reads of CTC and PBMC samples were trimmed using Trimmomatic tool (version 0.32) and then aligned to human reference genome (hg38) with STAR (version 2.6) (Dobin et al., 2013). Fragments Per Kilobase of exon per Million mapped fragments (FPKM) value was calculated with Cufflinks (version 2.2) (Trapnell et al., 2012). Read counts for each gene was determined using the HTSeq tool (version 0.11.2). Transcripts per million (TPM) values were determined using R as previously described (Li and Dewey, 2011). Counts per million (CPM) were estimated for each sample by using edgeR (Robinson et al., 2010).

Sample quality was assessed in the following steps: first, we determined sequencing quality by investigating the number of mapped reads and the number of expressed genes within each sample. Samples with less than 10 million reads were removed from the analysis. Second, unsupervised hierarchical clustering analysis and principal component analysis (PCA) were applied to confirm two distinct cell populations of CTCs and PBMCs. Third, to filter out CTC samples with leukocyte contamination, we adapted the experimental approach of Miyamoto *et al*., establishing filtering criteria for prostate cancer CTCs based on gene expression profiles (Miyamoto et al., 2015). A “Prostate cancer (PC) Signal” was determined for each sample by using the maximum FPKM value among PC markers (PSA, PSMA, AMACR and AR) or epithelial markers (KRT7, KRT8, KRT18, KRT19 and EPCAM). Similarly, a “Leukocyte Signal’’ was determined for each sample by taking the maximum value of the leukocyte markers CD45 and CD16. The median PC and Leukocyte Signals among all samples were taken as lower-bound thresholds for inclusion for further analysis.

### Differential gene expression (DGE) analysis and Gene sets enrichment analysis (GSEA)

Differential gene expression analysis was performed using DESeq2 (Love et al., 2014); resulting p-values were used to calculate False Discovery Rate (FDR) through Benjamini-Hochberg (BH) method. The threshold for differential expression used for all analyses was set at FDR ≤ 0.1 and fold change (FC)≥2. For the CTCs comparison with PBMCs, the threshold was FDR≤0.05 and FC≥2.

Enrichment of signaling pathways in the differentially expressed genes was determined by performing a pre-ranked gene set enrichment analysis (GSEA) using fgsea R package (Sergushichev, 2016) and the HALLMARK, Oncogenic and “Curated gene set” from MSigDB (Subramanian et al., 2005). P-values were determined using 1,000,000 permutations to estimate FDR using the BH method. We applied a stringent FDR threshold of 0.01 for all GSEA analysis with the exception of comparison between CTCs at PR with BS, for which we used FDR threshold of 0.25. Single sample enrichment scores of customized datasets for each patient were calculated using GSVA with the method of “ssgsea” (Hanzelmann et al., 2013) using genome-wide log2 TPM values as input.

### PBMC transcriptional analysis

PBMC transcriptomic profiles were analyzed using the xCell platform to evaluate abundance of 64 immune cell types (Aran et al., 2017). FPKM values were used as input to quantify the signature scores of each immune cell type at baseline and progression; Wilcoxon rank-sum test was used to statistically analyze the data. The resulting p-values were used to generate FDR values estimates using the BH method.

Immune modular analysis was also conducted using an established framework of 97 modules derived from whole blood transcriptomics (Banchereau et al., 2016; Chaussabel et al., 2008). The module score was calculated using the average log2 fold change of genes within a module after subtracting average gene expression of baseline PBMC. Differential module comparison analysis was conducted between 1) baseline and progression PBMC samples, and 2) baseline progressors and responders. Modules with p-value lower than 0.05 were considered significantly differentially enriched. Transcriptomic-based neutrophil-to-lymphocyte ratio (tr-NLR) score was calculated using neutrophil module (M5.15) and T cell (M4.1) ratio.

## List of Supplementary Materials

Table S1. Transcriptomic and genomic status of RB1, E2F1, BRCA1, BRCA2 and ATM genes in patients with mCRPC based on PFS classification.

Table S2. Proportional hazards regression analysis of transcript-based Neutrophil–Lymphocyte ratio (NLR) in baseline PBMC predicts overall survival (OS) and progression-free survival in patients with mCRPC.

Fig. S1. Reads and gene number distribution in all samples.

Fig. S2. Number of mapped and assigned reads across CTC and PBMC samples obtained at baseline (BS, n=40) and at progression (PR, n=22).

Fig. S3. CTC samples were candidated based on prostate-lineage and epithelial-lineage marker expression.

Fig. S4. CTCs and PBMCs are two distinct cell populations based on principal component analysis (PCA) and differential gene expression analysis.

Fig. S5. Kaplan Meier Survival Curve of RB loss signature and E2F target gene signatures. Fig. S6. Receiver operating characteristic (ROC) curve of RB and E2F signature scores.

Fig. S7. BRCA40 Core Genes of BRCA networks and their correlations with E2F and RB signatures.

Fig. S8. Deconvolution xCell analysis of PBMC transcriptomic profiles identifies total macrophage enrichment and CD8+ T-cells depletion at progression (PR) relative to baseline (BS).

Fig. S9. Heatmap of gene expression values of differentially expressed modules between baseline (BS) and progression (PR).

Fig. S10. Enrichment of myeloid lineage, inflammation and neutrophil modules with concomitant depletion of T cell and cytotoxicity NK cell modules at progression (PR) was confirmed using GSEA.

## Data Availability

All data produced in the present study are available upon reasonable request to the authors

## Acknowledgments

We wish to thank the Prostate Cancer Foundation and Movember for their financial support of this Global Treatment Sciences Challenge Award, and the US Department of Defense Prostate Cancer Clinical Trial Consortium for infrastructural support for this multicenter study. Dr. Giannakakou was supported by the Department of Defense grant W81XWH-18- PCRP-IDA and by the National Cancer Institute of the National Institutes of Health under Award Number T32CA203702 (to J.Zhang), by T32CA062948 (to G.Galletti) by R01 CA137020 (to P. Giannakakou), by R01 CA179100 (to P. Giannakakou) by R21 CA216800 (to P. Giannakakou). Dr. Armstrong was supported by a Prostate Cancer Foundation grant, an NIH R01 (1R01CA233585-01) and the DCI P30 CA014236 as well as Duke Cancer Institute shared resources for biostatistics, Flow Cytometry, and Sequencing and Genomic Technologies. This work was partially funded by Department of Defense grants W81XWH- 13-PCRP-CCA, W81XWH-17-2-0021 and W81XWH-14-2-0179 (DJG/AJA, Duke), W81XWH-15-1-0467 (SH, Duke), W81XWH-18-1-0278 (SH, Duke), W81XWH-14- 2-0159 (DMN, Weill Cornell), W81XWH-15-2-0018 (RS, Chicago), W81XWH-15-2-0018 (HIS, MSKCC), and W81XWH-16-PCRP-CCRSA (E.S.A, Johns Hopkins). This research was supported in part by National Cancer Institute (NCI) Grant NIH T32CA062948 and the CTSC UL1 TR002384-01 and P30 CA014236. We wish to thank the study coordinators at Weill Cornell, Duke University, Johns Hopkins, University of Chicago, and Memorial Sloan Kettering Cancer Center. We wish to acknowledge the dedication of our patients to provide blood samples at no clear benefit to them but for the benefits of all patients with prostate cancer.

## Author contributions

Funding acquisition: A.J.A and P.G. Study design: P.G. Clinical patients’ blood collection: G.G., S.G., M.A., D.G., S.G., D.N., S.T. and A.J.A. Sample processing for RNA sequencing: J.Z., G.G., and A.G. Bioinformatic investigation of CTCs: J.Z., B.Z., A.V., A.S., and O.E. PBMC derived modular analysis: J.Z., B.Z., S.H., V.P. and C.M. Original draft: J.Z., G.G., and P.G. All authors have reviewed the data analyses, contributed to intellectual content of the manuscript and approved the final version of manuscript to be published.

