## Supplemental file for "Liquid-biopsy transcriptomic profiling uncovers molecular mediators of resistance to androgen receptor signaling inhibition in lethal prostate cancer"

**Supplementary Materials**

**Table S1. Transcriptomic and genomic status of RB1, E2F1, BRCA1, BRCA2 and ATM genes in patients with mCRPC based on PFS classification.**

|  | RB/E2F common signature transcriptomic status | E2F targets signature transcriptomic status | E2F1 genomic status (CNV/Mutation) | RB loss signature transcriptomic status | RB1 genomic status (CNV/Mutation) | BRCA40 signature transcriptomic status | BRCA1 genomic status (CNV/Mutation) | BRCA2 genomic status (CNV/Mutation) | ATM genomic status (CNV/Mutation) |
| --- | --- | --- | --- | --- | --- | --- | --- | --- | --- |
| Progressors, PFS ≤ 3 months | High | Low | Neutral/WT | High | Amplification/WT | Low | Amplification/Missense | Neutral/Missense | Neutral/WT |
|  | High | High | Neutral/WT | High | Neutral/WT | High | Neutral/WT | Neutral/WT | Neutral/WT |
|  | High | High | Neutral/WT | High | Neutral/WT | High | Neutral/WT | Neutral/WT | Neutral/WT |
|  | High | High | Neutral/WT | High | Neutral/Missense | Low | Neutral/Missense | Neutral/Missense | Neutral/Missense |
|  | Low | Low | Neutral/WT | Low | Neutral/Missense | Low | Neutral/Missense | Neutral/Missense | Neutral/Missense |
|  | High | Low | Neutral/WT | High | Neutral/WT | Low | Neutral/WT | Neutral/WT | Neutral/WT |
|  | Low | Low | Neutral/WT | Low | Amplification/ | Low | Amplification/ | Neutral/WT | Neutral/WT |
|  | Low | Low | Neutral/WT | Low | Neutral/WT | Low | Neutral/WT | Neutral/WT | Neutral/WT |
|  | Low | Low | NA | Low | NA | Low | NA | NA | Neutral/WT |
|  | Low | Low | Neutral/WT | Low | Neutral/WT | Low | Neutral/WT | Neutral/WT | Neutral/WT |
| Patients with PFS > 3 and < 12 months | High | High | Neutral/WT | High | Neutral/WT | High | Neutral/WT | Neutral/WT | Amplification/WT |
|  | Low | Low | Neutral/WT | Low | Neutral/WT | Low | Neutral/WT | Neutral/WT | Neutral/WT |
|  | Low | Low | Neutral/WT | High | Neutral/WT | Low | Neutral/WT | Neutral/WT | Neutral/WT |
|  | Low | Low | NA | Low | NA | Low | NA | NA | Neutral/WT |
|  | Low | Low | loss/WT | Low | Neutral/WT | Low | Loss/Missense | Neutral/Missense | Neutral/Missense |
|  | High | Low | Neutral/WT | High | Neutral/WT | High | Neutral/WT | Neutral/WT | Amplification/WT |
|  | Low | Low | Neutral/WT | Low | Neutral/WT | Low | Neutral/WT | Neutral/WT | Neutral/WT |
|  | Low | Low | Neutral/WT | High | Neutral/WT | High | Neutral/WT | Neutral/WT | Neutral/WT |
|  | Low | Low | Neutral/WT | Low | Neutral/Missense | Low | Neutral/Missense | Neutral/Missense | Neutral/Missense |
|  | Low | Low | Neutral/WT | Low | Neutral/WT | Low | Neutral/WT | Neutral/WT | Neutral/WT |
|  | High | High | Neutral/WT | High | Neutral/WT | High | Neutral/WT | Neutral/WT | Neutral/WT |
| Responders, PFS ≥ 12 months | High | High | Neutral/NA | High | Neutral/WT | High | Neutral/WT | Neutral/WT | Neutral/WT |
|  | High | High | NA | High | NA | Low | NA | NA | Neutral/WT |
|  | Low | Low | Neutral/NA | Low | Neutral/WT | Low | Neutral/WT | Neutral/WT | Amplification/WT |
|  | Low | Low | NA | Low | NA | Low | NA | NA | Neutral/WT |
|  | Low | Low | Neutral/NA | Low | Neutral/WT | Low | Neutral/WT | Neutral/WT | Neutral/WT |
|  | Low | Low | Neutral/NA | Low | Neutral/WT | Low | Neutral/WT | Neutral/WT | Neutral/WT |
|  | Low | Low | Neutral/NA | Low | Neutral/WT | Low | Neutral/WT | Neutral/WT | Neutral/WT |
|  | Low | Low | Neutral/NA | Low | Neutral/WT | Low | Amplification/WT | Neutral/WT | Neutral/WT |

Note: WT represent Wild type; NA means Not available

**Table S2. Proportional hazards regression analysis of transcript-based Neutrophil–Lymphocyte ratio (NLR) in baseline PBMC predicts overall survival (OS) and progression-free survival in patients with mCRPC.**

|  | NLR | | | P value |
| --- | --- | --- | --- | --- |
|  | Low (<= 1.143) | High (>1.143) | |  |
| Median OS (months; 95% CI) | 29.2 (22.4-NR) | 15.9 (8.2-27.3) | |  |
| HR (95% CI), dichotomous NLR | 2.8 (1.3-5.8) | | | 0.008 |
| HR (95% CI),  continuous NLR | 4.9 (1.4-17.3) | | | 0.014 |
| Median PFS (months; 95% CI) | 8.4 (3.1-21.5) | | 5.9 (3.5-13) |  |
| HR (95% CI), dichotomous NLR | 1.7 (0.9-3.3) | | | 0.14 |
| HR (95% CI),  continuous NLR | 3.0 (0.9-9.5) | | | 0.065 |

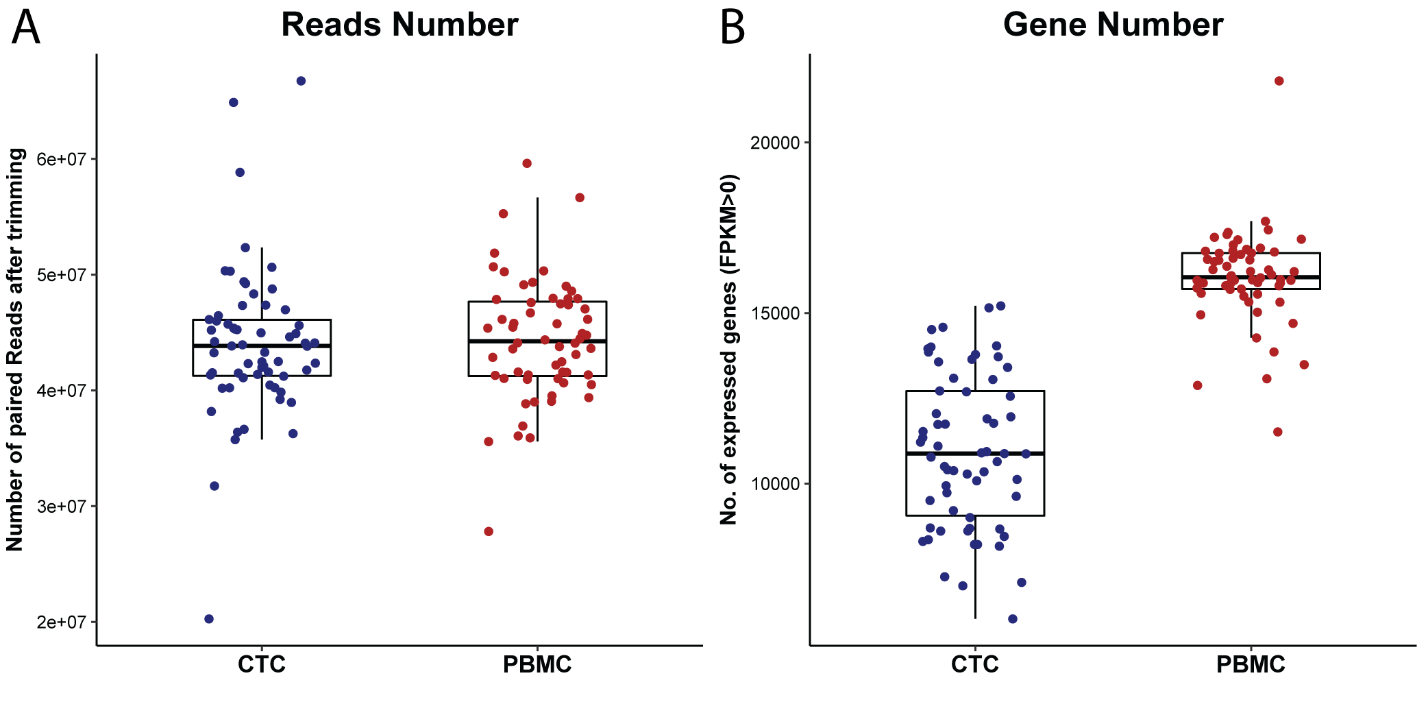

**Fig. S1. Reads and Gene number distribution in all samples. A.** After trimming raw reads, an average of 44.0 million (range from 20.3 million to 66.8 million) paired reads per CTC sample and 44.3 million (range from 27.8 million to 59.6 million) paired reads for PBMCs sample remained, indicating consistent sequencing yield across samples. **B.** We detected expression of 10,947 genes on average in CTCs, while in PBMCs there were 16,045 genes expressed, defined as having at least one read mapped (Figure S1B). The 5,000 genes difference between CTCs and PBMCs suggested a higher expression complexity in PBMC samples, which could be attributed to the greater diversity of cell types in PBMCs.

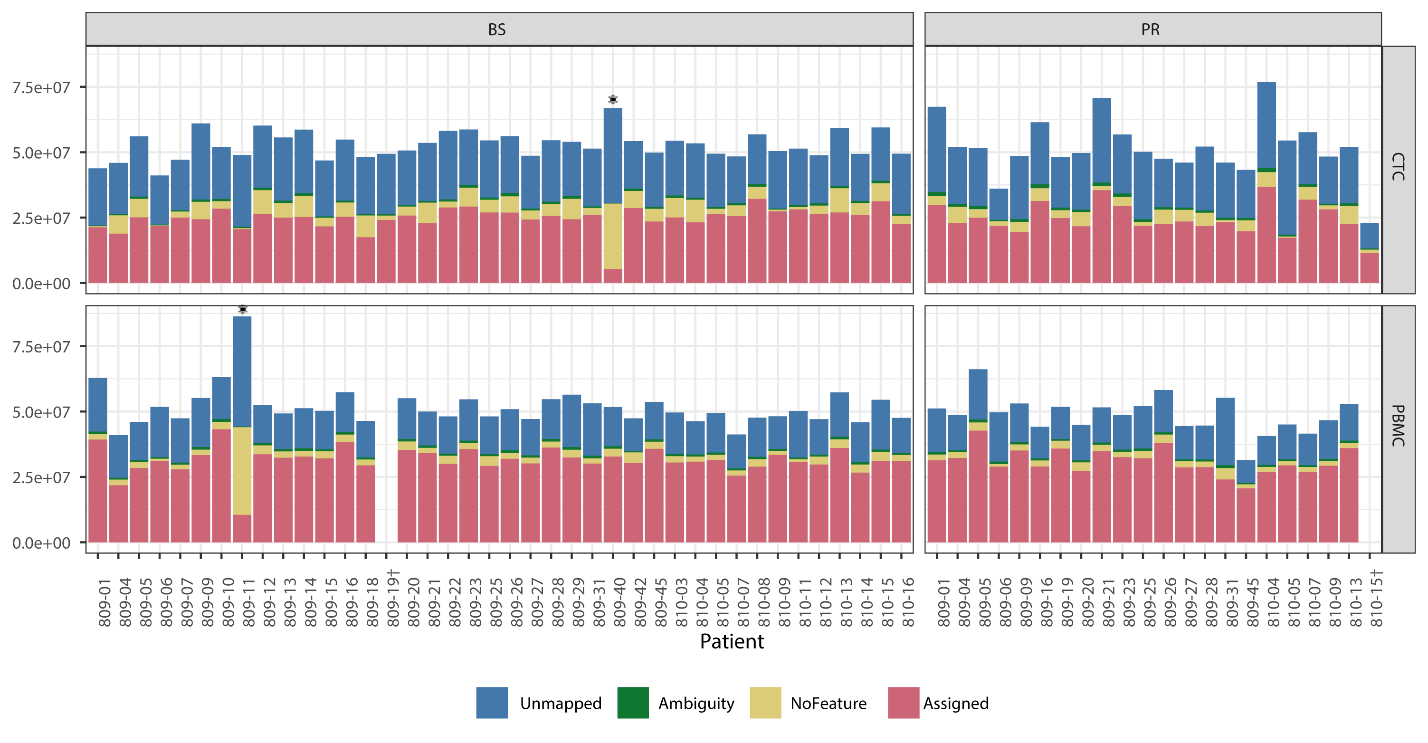

**Fig. S2. Number of mapped and assigned reads across CTC and PBMC samples obtained at baseline (BS, n=40) and at progression (PR, n=22)**. * Samples with fewer than 10,000,000 assigned reads were filtered out. † represents missing PBMC samples during processing.

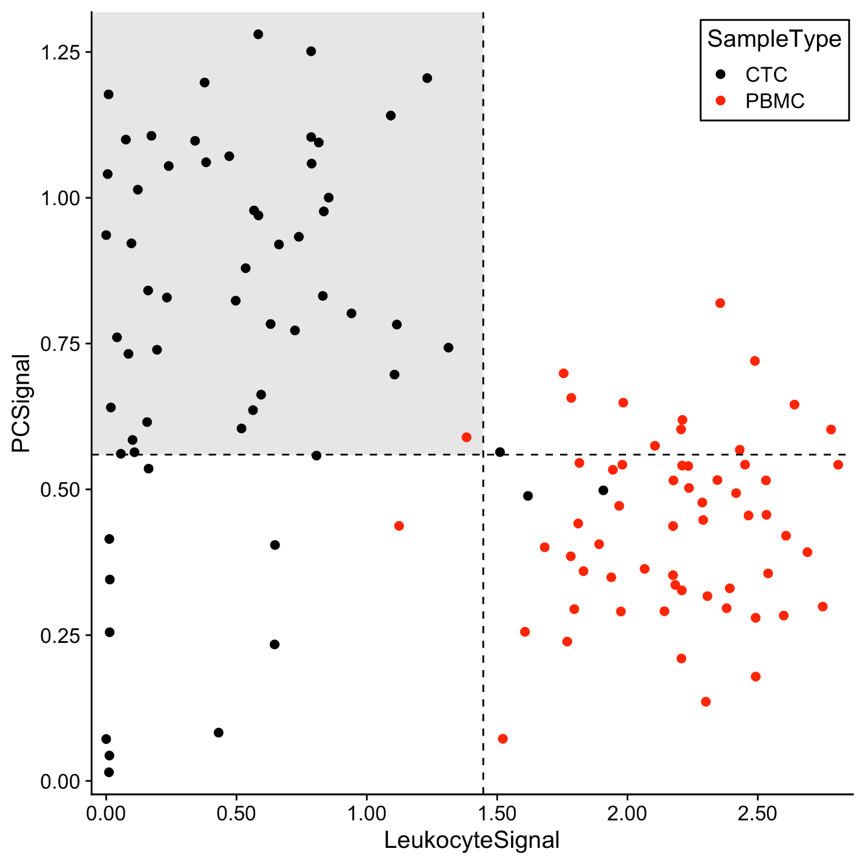

**Fig. S3. CTC samples were candidated based on prostate-lineage and epithelial-lineage marker expression.** Strict expression thresholds were used to define lineage-confirmed CTCs, scored by prostate lineage-specific transcripts (PSA, PSMA, AMACR, and AR) and standard epithelial markers (KRT7, KRT8, KRT18, KRT19, and EPCAM) adapted from Miyamoto et al., 2015. PBMCs were distinguished by expression of the hematopoietic lineage markers CD45 and CD16 and served to exclude any CTCs with potentially contaminating signals. Dotted lines represent the median signal of all the samples. Given the presence of leukocyte transcripts suggestive of cellular contamination or misidentification during selection, 3 CTC samples were excluded, and given low expression of both prostate lineage-specific transcripts and standard epithelial markers, 11 CTC samples were further excluded. The remaining 47 CTC samples (shaded area in the plot) included 29 BS and 18 PR samples were used for all downstream analyses. Similarly, 2 PBMC samples were removed based on low expression of leukocytes transcripts leaving 57 evaluable samples, including 37 at BS and 20 at PR.

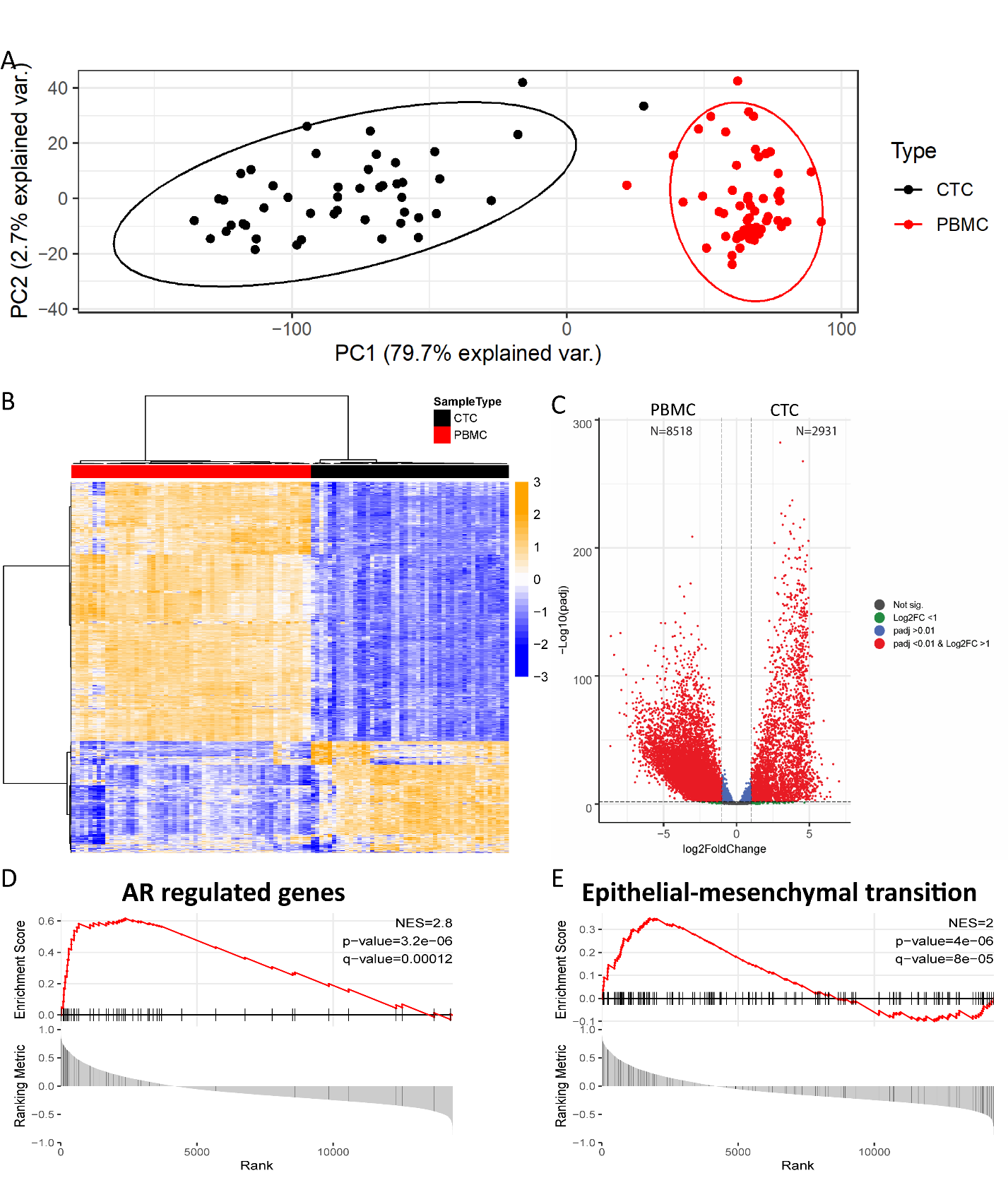

**Fig. S4 CTCs and PBMCs are two distinct cell populations based on principal component analysis (PCA) and differential gene expression analysis. S4A.** CTCs and PBMCs are clearly separated by the first Principal component (PC1) of gene counts**.** Transcriptional profiles of CTC and PBMC patient samples embedded into the first two principal components using the 500 genes with the highest variance. Lines represent the 95% confidence ellipsoids of each sample type. The first principal component (PC1) explains 79.7% of the variance, confirming that the vast majority of the variance derives from the difference between the two sample types, ruling out confounding factors such as batch effects or patient of origin. **S4B.** Unsupervised hierarchical clustering analysis identified CTCs and PBMCs as two distinct cell clusters. Heatmap shows the top 2,000 differentially expressed genes selected based on threshold of q-value <0.01 and fold change >2. **S4C**. Volcano plot analysis further shows 8,518 and 2,931 genes differentially expressed in PBMCs and CTCs, respectively. **S4D-E**. Gene Set Enrichment Analysis (GSEA) using the Molecular Signatures Database (MSigDB) identified the Epithelial-mesenchymal transition (EMT) pathway and the AR activated genes pathway (REACTOME_ACTIVATED_PKN1_STIMULATES_TRANSCRIPTION_OF_ANDROGEN_RECEPTOR_REGULATED_GENES_KLK2_AND_KLK3) as significantly enriched in CTCs.

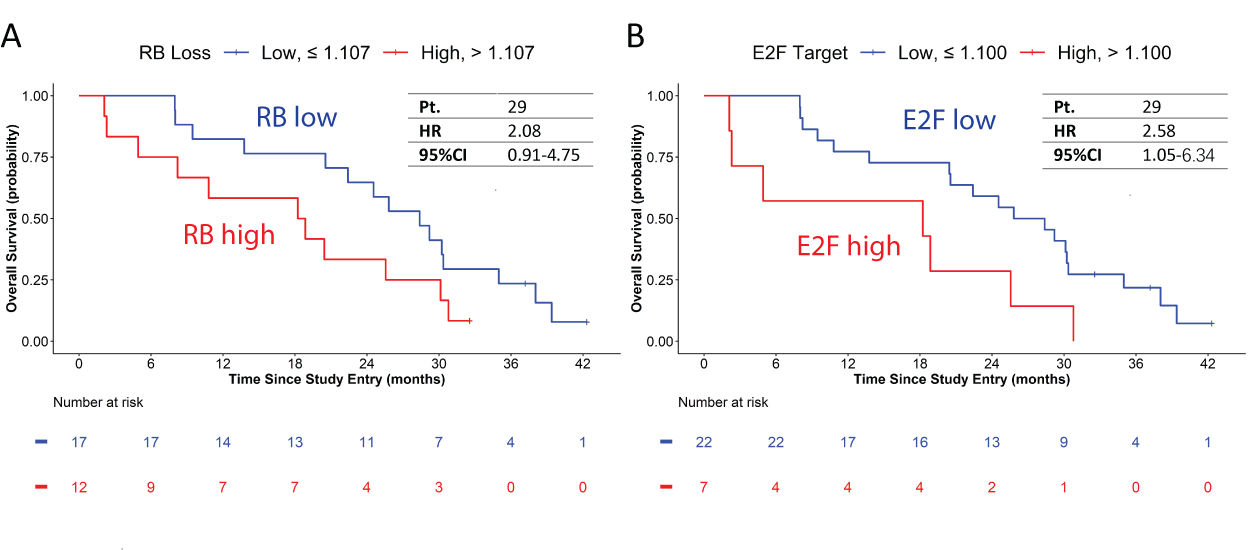

**Fig. S5. Kaplan Meier Survival Curve of RB loss signature and E2F target gene signatures. S5A.** Kaplan-Meier curves displaying the estimated survival probability for 2 groups of mCRPC patients at baseline (BS) with optimized dichotomous cutoff of high or low RB score. The optimal dichotomous cutoff of high or low RB score was determined by assessing the association between RB score and overall survival using maximally selected rank statistics (Lausen and Schumacher, 1992) Overall survival was evaluated using a univariate proportional hazards (CoxPH) model, and hazard ratio (HR) and range were shown in the table. Vertical mark in the curve indicates censored patients. **S5B.** Kaplan-Meier curves displaying overall survival probability for 2 different groups of patients with mCRPC at baseline (BS) with either high or low E2F score. Overall survival survival by RB loss and E2F were evaluated using a univariate proportional hazards (CoxPH) model. Hazard ratio (HR) and confidence interval (CI) were shown in the table. Vertical mark in the curve indicates censored patients.

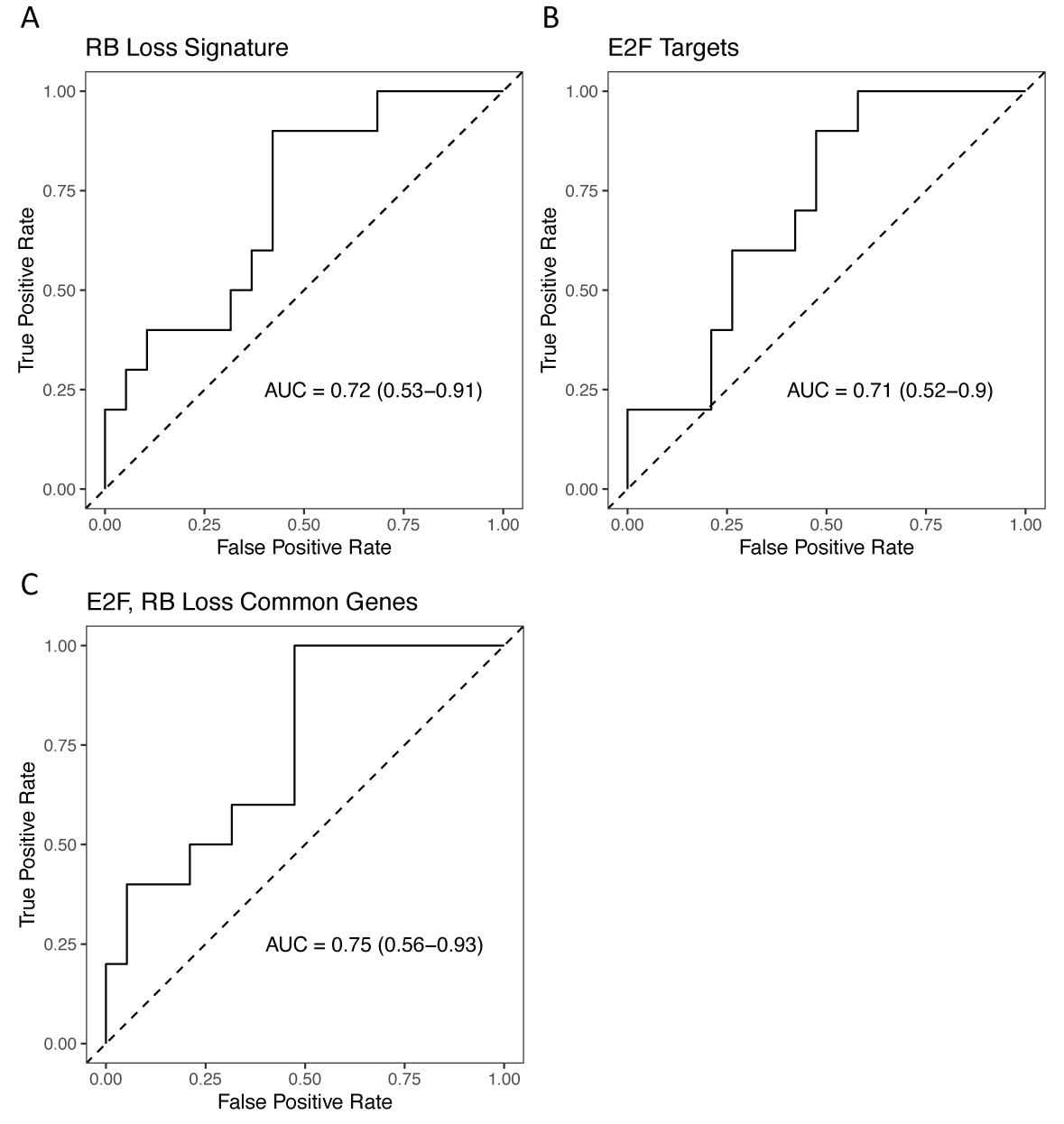

**Fig. S6. Receiver operating characteristic (ROC) curve of RB and E2F signature scores**. Single sample enrichment score (ssGSEA) was calculated through GSVA using log2 TPM values as input to classify Progressors from the cohort. 95% confidence intervals, calculated using the DeLong method, are shown in parentheses. **S6A**. RB loss signature score ROC curve. RB loss signature (Ertel et al., 2010, n=158 genes) gene set. **S6B**. E2F targets signature (MSigDB, n=200 genes) score ROC curve. **S6C**. RB/E2F common signature score ROC curve.

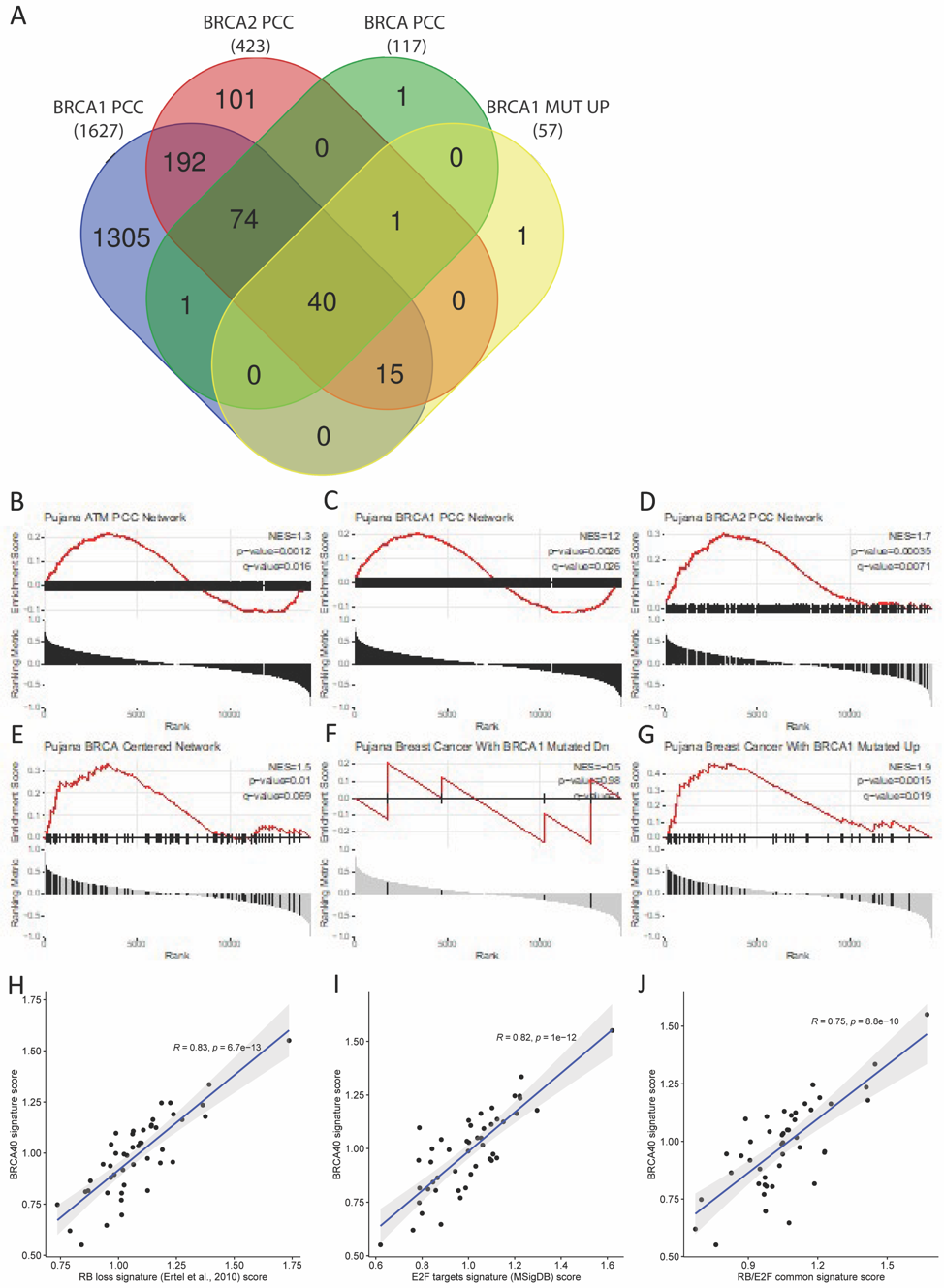
**Fig. S7. BRCA40 Core Genes of BRCA networks and their correlations with E2F and RB signatures. S7A.** Venn diagram of BRCA1 network, BRCA2 network, BRCA network, and BRCA1 KO up-regulated gene signatures (Pujana et al., 2007). **S7B-G.** Enrichment plots of BRCA-related gene sets, identified in breast cancer (Pujana et al 2007), in BRCA-altered SU2C patients as compared to unaltered patients. **S7H-J** BRCA40 signature correlates highly and significantly with RB loss (Ertel et al, 2010), E2F target genes (MsigDB) and the RB/E2F common signature described here, in all CTC samples.

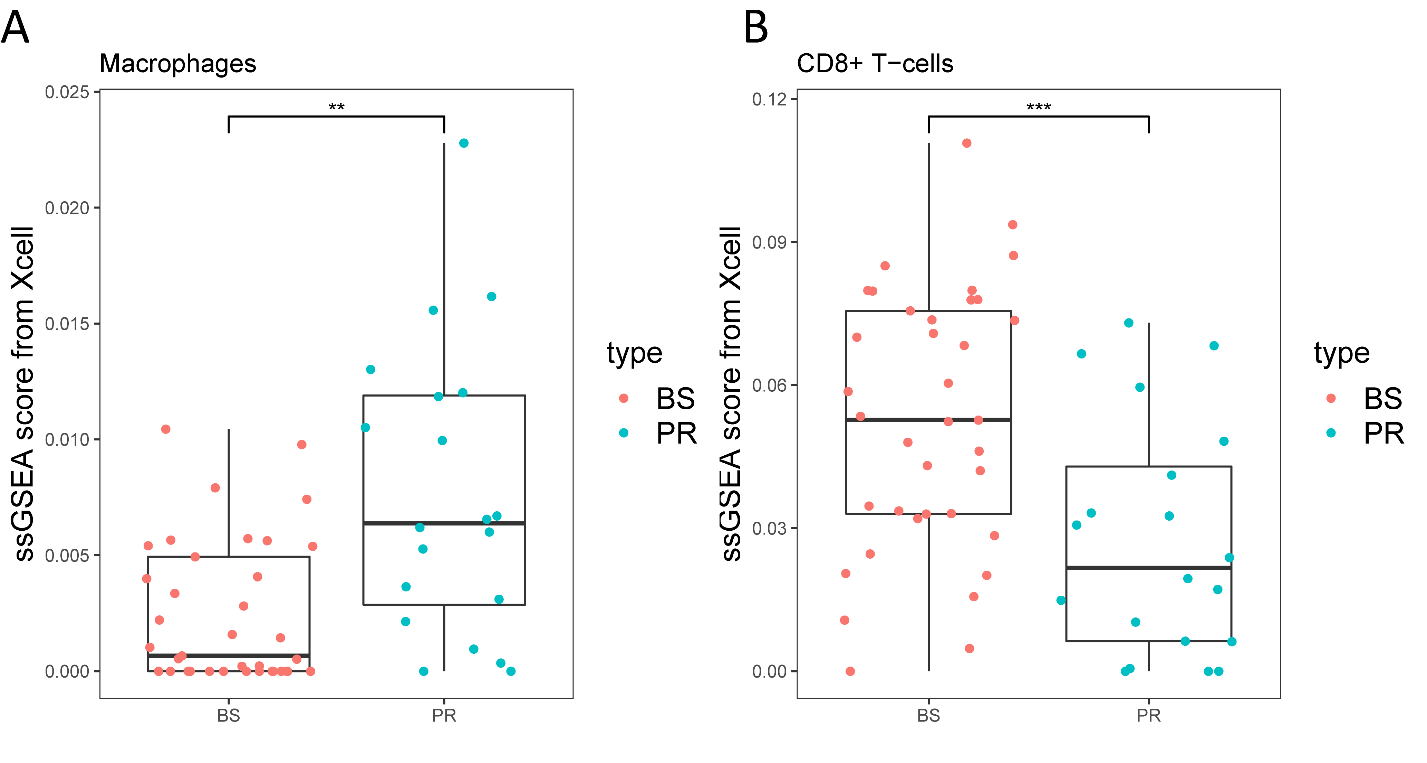

**Fig. S8. Deconvolution xCell analysis of PBMC transcriptomic profiles identifies total macrophage enrichment and CD8+ T-cells depletion at progression (PR) relative to baseline (BS).** Cell type enrichment analysis was performed using FPKM values as input for 64 immune and stroma cell types. Statistical significance was determined by Wilcoxon t test using ssGSEA score. * p-value <0.01; ** p-value < 0.001. Macrophage up regulation (**S8A**) and CD8+ T-cells down-regulation (**S8B**) was in PR.

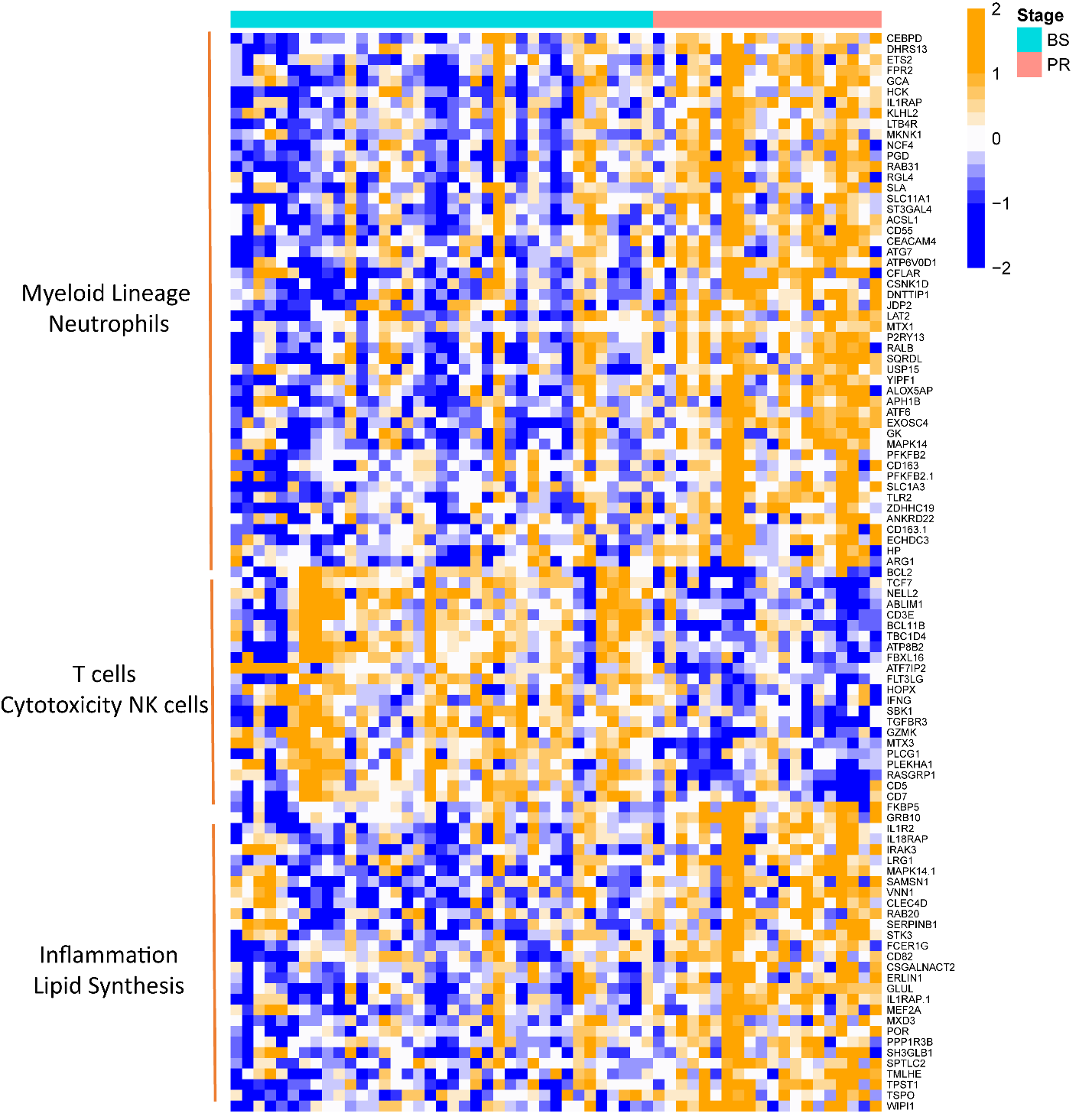

**Fig. S9. Heatmap of gene expression values of differentially expressed modules between baseline (BS) and progression (PR).** Myeloid Lineage, Neutrophils, Inflammation and Lipid Synthesis modules were up-regulated in Progression PBMCs, while T-cells and Cytotoxicity NK cells are down-regulated in PR PBMCs compared with BS PBMCs. Figure shows supervised heatmap of 101 significant genes in 10 significant immune modules from the comparison of PR PBMCs with BS PBMCs. P adjust value cutoff is 0.05. Heatmap columns were ordered by their patient ID and time point of blood collection.

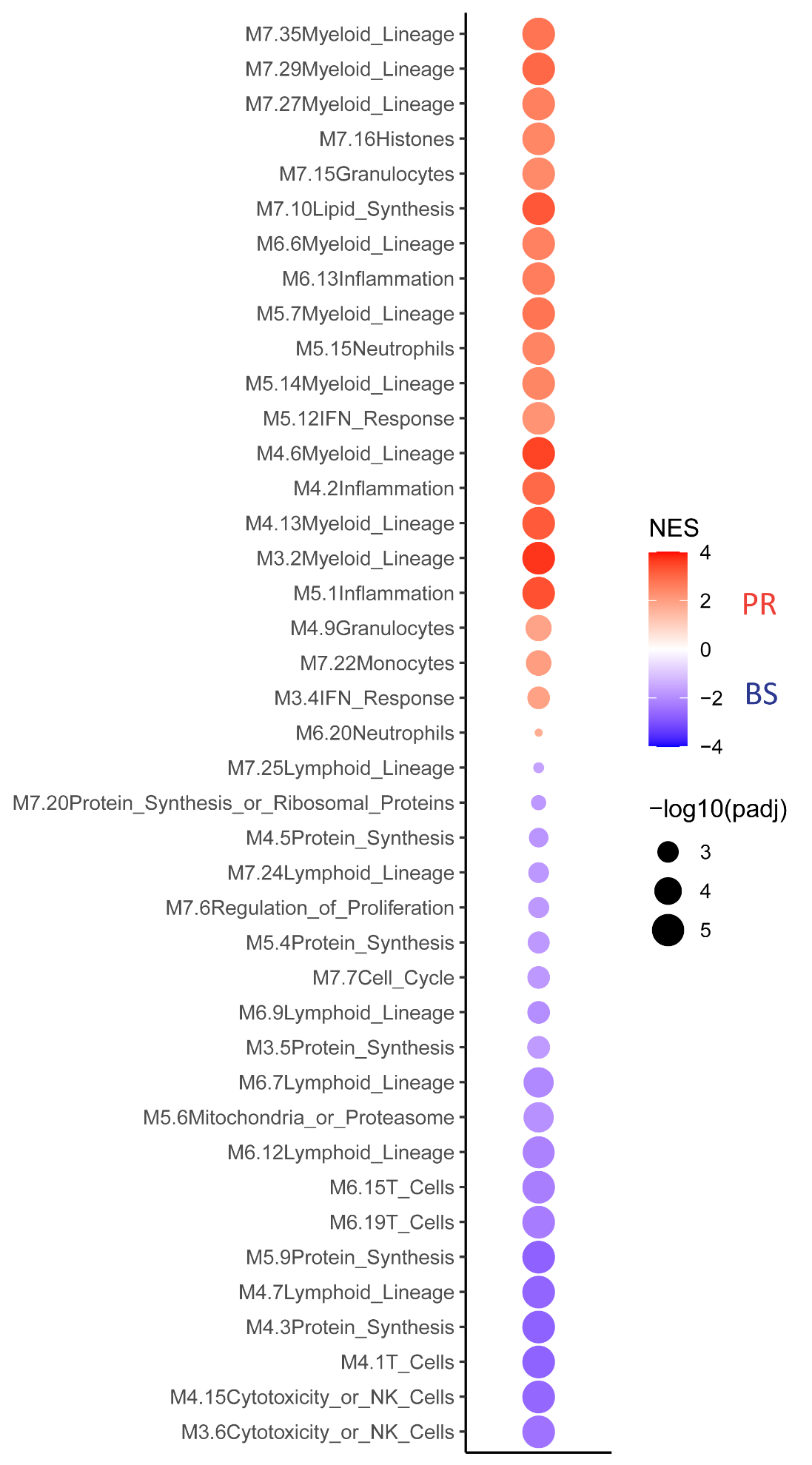

**Fig. S10. Enrichment of myeloid lineage, inflammation and neutrophil modules with concomitant depletion of T cell and cytotoxicity NK cell modules at progression (PR) was confirmed using GSEA.** Of the 97 immune modules described (in Chaussabel et al., 2008; Banchereau et al., 2016), the 63 which have a functional annotation were used as a customized database for GSEA analysis. The p-value cutoff is 0.01.
